# Cognitive subgroups of affective and non-affective psychosis show differences in medication and cortico-subcortical brain networks

**DOI:** 10.1101/2022.08.03.22278370

**Authors:** Katharina M Bracher, Afra Wohlschlaeger, Kathrin Koch, Franziska Knolle

**Affiliations:** Division of Neurobiology, Faculty of Biology, LMU Munich, Martinsried 82152, Germany; Department of Diagnostic and Interventional Neuroradiology, School of Medicine, Technical University of Munich, Munich, Germany

**Author notes:** Author contributions: FK, KB, AW conceptualisation; KK preprocessing of structural brain data; FK and KB formal analysis of cognitive data and clustering analysis; FK and KB writing of first draft; FK, KB, AW and KK editing and revising of manuscript. Data availability statement: All processed data and code may be accessed via: https://github.com/KatharinaBracher/earlypsychosis_clustering.

## Abstract

Cognitive deficits are prevalent in individuals with psychosis and are associated with neurobiological changes, potentially serving as an endophenotype for psychosis. Using the HCP-Early-Psychosis-dataset (n=226), we aimed to investigate cognitive subtypes (deficit/intermediate/spared) through data-driven clustering in affective (AP) and non-affective psychosis patients (NAP) and controls (HC). We explored differences between three clusters in symptoms, cognition, medication, and grey matter volume. Applying principal component analysis, we selected features for clustering. Features that explained most variance were scores for intelligence, verbal recognition and comprehension, auditory attention, working memory, reasoning and executive functioning. Fuzzy K-Means clustering on those features revealed that the subgroups significantly varied in cognitive impairment, clinical symptoms, and, importantly, also in medication and grey matter volume in fronto-parietal and subcortical networks. The spared cluster (86%HC, 37%AP, 17%NAP) exhibited unimpaired cognition, lowest symptoms/medication, and grey matter comparable to controls. The deficit cluster (4%HC, 10%AP, 47%NAP) had impairments across all domains, highest symptoms scores/medication dosage, and pronounced grey matter alterations. The intermediate deficit cluster (11%HC, 54%AP, 36%NAP) showed fewer deficits than the second cluster, but similar symptoms/medication/grey matter to the spared cluster. Controlling for medication, cognitive scores correlated with grey matter changes and negative symptoms across all patients. Our findings generally emphasize the interplay between cognition, brain structure, symptoms, and medication in AP and NAP, and specifically suggest a possible mediating role of cognition, highlighting the potential of screening cognitive changes to aid tailoring treatments and interventions.

## 1 Introduction

Cognitive alterations are core symptoms of psychosis [1–4], which have been described in areas of working memory [5, 6], attention [7, 8], reasoning [9, 10], decision-making [11– 13], salience processing [14, 15], learning (e.g., lower learning rate or increased forgetting, [16–18]) and problem solving [19, 20] and across all stages of the disease [21–25]. Cognitive deficits furthermore precede the clinical onset of psychosis [26], and predict functional outcome in later stages of the disease [27, 28], impacting employment status, independent living and social functioning [28, 29]. Although cognitive impairments are present in about 80% of patients suffering from psychotic disorders [30–32], therapeutic interventions are limited. A meta-analysis including 93 studies using different agents targeting mainly glutamatergic and cholinergic neurotransmitter systems, but also serotonin, dopamine, GABA and noradrenaline agents [33] reported a significant, although very small (g=0.10) improvement of cognition in general. However, this meta-analysis failed to find significant improvement for cognitive subdomains [33]. Cognitive training, also called cognitive remediation therapy, has produced more promising effects [34, 35]. A recent meta-analysis found that cognitive remediation showed significant small-to-moderate cognitive improvements in all domains studied (g=.19–.33) and a small improvement in function (g=.21). Furthermore, research has shown that fewer cognitive deficits and higher cognitive reserve during prodromal and first episode psychosis are generally, independent of diagnosis, associated with better functioning and recovery [36, 37]. This indicates that maintaining and improving cognitive functioning is crucial in the interventional and therapeutic processes.

Cognitive deficits in psychosis, in general, have been linked to alterations in the corticocerebellar-thalamic-cortical circuits [38]. Here, dysfunctional GABA (gamma-amino-butyric acid) inter-neurons, the main inhibitory neurons of the central nervous system, may disrupt the balance between excitatory and inhibitory processes in the cortex [39]. A recent review [25] summarizes the association between functional brain alterations and cognitive deficits in individuals at-risk for psychosis, early onset psychosis, and chronic schizophrenia. They report a clear association between altered cortical (e.g., prefrontal cortex, anterior cingulate cortex, insula) and subcortical (e.g., thalamus, striatum, hippocampus, cerebellum) brain signalling and aberrant cognition across the different stages. Imaging studies provide further support showing reduced grey matter volume and altered network organization which correlates with cognitive deficits at illness onset [40, 41], early psychosis [42] and chronic schizophrenia [43]. Furthermore, a study by Van Rheenen et al. [44] reported global grey matter volume and thickness reductions which were more prominent in patients with stronger cognitive impairments.

Interestingly, psychosis patients with different diagnoses, e.g., affective vs non-affective psychosis, show varying cognitive deficits [45–47]. In a review, Barch and Sheffield [47] summarized that while the severity of cognitive impairment is stronger in non-affective compared to affective psychosis, the relative impairments across different cognitive domains are very similar. Other studies, however, do not differentiate between affective vs non-affective psychosis when investigating cognitive deficits [34, 48, 49]. Despite these differences in cognitive alterations with regard to specific diagnoses, cognitive deficits in psychosis have been described and investigated as intermediate phenotypes [50]. In a recent study, Shafee et al. [51] revealed that cognitive phenotypes may vary grossly depending on specific types of psychosis (e.g. affective vs non-affective), suggesting that certain domains of cognition (e.g., working memory vs face processing) may be more etiologically linked to psychosis than others. Using a K-means clustering approach in a cross-diagnostic sample, Lewandowski et al. [52] identified four cognitive subgroups combining different psychosis groups. Importantly, they identified one cognitively intact cluster consisting of healthy controls and patients with different diagnoses, while the other three clusters were dominated by different cognitive impairment profiles [52]. The data-driven identification of cognitive clusters varies between two and four-cluster solutions, as shown in a recent systematic review [53]. One major contributing factor may be the variance in the data, especially the cognitive measures that are being used. Interestingly, a recent systematic review of data-driven identification of cognitive subtypes [54] highlighted that despite the heterogeneity of clustering methods used and cognitive domains studied, there is some commonality in the identification of a severe cognitive deficit phenotype showing deficits across multiple domains, and a spared cognitive deficit phenotype with similar performances to controls. While it is unclear from the literature which cluster solution is the most reliable, especially an intermediate phenotype may be interesting from an interventional point of view, as this group of individuals may benefit the most from cognitive interventions to improve functional outcome. It has been shown [55] that patients at early compared to late illness stages have a higher potential for cognitive improvement after cognitive remediation therapy. This may be directly translatable to an intermediate cognitive phenotype, as this group may possess residual cognitive abilities that can be enhanced through targeted interventions and may also have more intact neural substrates that support cognitive training. Furthermore, those cognitive improvements may be more likely to translate into functional gains, such as better social and occupational outcomes, which are crucial for improving the overall quality of life [56–58]. Additionally, all studies using cognitive clustering concentrated on cognitive toolboxes, or basic measures such as measures of processing speed or working memory. Using simple task data, however, such as the Delay Discounting task may be a beneficial contribution as they include reward processing an ability reported to be impaired across all stages of psychosis [12, 13, 15, 17]. Furthermore, it is unclear how cognitive subtypes are linked to differences in medication status and brain structure (i.e., grey matter volume). These open questions, however, are crucial to understand whether cognitive subtypes are clinically relevant, and whether they could increase our mechanistic understanding of the disorder.

In the current study, we, therefore, aimed to explore these important questions. First, we wished to investigate three cognitive subtypes using the HCP Early Psychosis dataset (https://www.humanconnectome.org/study/human-connectome-project-for-early-psychosis, [59]) using data-driven clustering on standardized cognitive, perceptual and emotional task and score data, but no clinical data. Second, we explored whether the patients in the three clusters differed in symptom expression, cognition, medication and grey matter volume. And third, depending on the results for the first two questions, we wished to understand if symptoms, alterations in cognition and brain morphometry were linked while controlling for medication within and across the clusters.

## 2 Methods

### 2.1 Participants

We analyzed data collected by the “Human Connectome Project for Early Psychosis” (HCP-EP, [60], [49]). The HCP-EP 1.1 release (August 2021 HCP-EP Release 1.1 on NDA) contains 251 subjects consisting of 68 healthy control individuals, 57 patients with affective and 126 patients with non-affective psychosis, both patient groups were within the first three years of the onset of psychotic symptoms. The Structured Clinical Interview for DSM-5: Research Version (SCID-5-RV) [61] was used to confirm diagnoses of non-affective (i.e., schizophrenia, schizophreniform, schizoaffective, psychosis not otherwise specified, delusional disorder, brief psychotic disorder) or affective psychosis (i.e., major depression with psychosis or bipolar disorder with psychosis). Clinical symptoms were assessed using the Positive and Negative Syndrome Scale (PANSS, [62]). Disease onset for all patients was within the last five years prior to study enrollment. For a comprehensive cognitive, perceptual and emotional assessment, the NIH toolbox ([63], [64]; i.e., cognition (Picture Sequence test, Dimensional Change test, Flanker test, Picture Vocabulary test, Pattern Completion test, List Scoring test, and Oral Reading test), emotion (Self-report emotion), perception (Words in Noise, Odor Identification, and Dynamic Visual Acuity), sensory-motor functions (9-Hole Pegboard, and Grip Strength), the HCP Lifespan Measures ([65]; i.e., Delay Discounting and Penn Emotion Recognition), the WASI-II [66] and the Seidman Auditory Continuous Performance Test [67] was used. Structural brain imaging data was available for 183 of the 251 subjects. After ensuring that there were sufficient data for features and subjects (see description of analysis below), the analysis was performed on 226 subjects (i.e., 56 healthy controls, 52 affective psychosis group, 118 non-affective psychosis group). Demographics and clinical scores for the three groups are presented in Tab. 1. Detailed analysis of PANSS items are presented in Suppl. Fig. 1.

### 2.2 Ethics statement

All participants provided written informed consent to participate in the HCP-EP, and for their data to be shared openly. HCP-EP was reviewed and approved by the Human Connectome Project (HCP). The HCP-EP complied with the ethical standards of the relevant national and institutional committees on human experimentation and with the code of ethics of the World Medical Association, the Helsinki Declaration of 1975, as revised in 2013.

### 2.3 Variable selection and preprocessing

Initially, all cognitive, perceptual and social/emotional functioning scores available in the HCP-EP dataset were selected, resulting in 70 variables as potentially relevant to our analysis (Suppl. Tab. 1). As covariates, we chose age, gender, socio-economic status and mother’s level of education. Variables containing information of primary diagnosis for affective and non-affective psychosis, such as the PANSS or the Clinical Assessment Interview for Negative Symptoms, as well as variables describing medication dosage, usage or equivalent doses, were not included in the clustering analysis, but only used for subsequent analyses.

Many of the selected variables contained missing data. We therefore excluded variables when more than 10% of the entries were missing, and we excluded subjects with more than 20% missing variables (Suppl. Tab. 1). As a result, 33 out of the initial 70 variables remained in the data set and 226 subjects. The distribution of the remaining subjects reflected the distribution of the original data with regard to diagnosis type. For an overview of selected variables and group comparison, see Suppl. Tab. 2. In the final dataset used for analysis, subjects were missing a maximum of seven variables (Suppl. Tab. 3 and Missing data were imputed using the mean for continuous and mode for categorical variables (e.g. demographic control variables). Since the ratio of patients and controls was not balanced, mean or mode for data imputation was calculated separately for patients and controls. Both patient groups were combined in order to minimize the bias of classical group membership (Suppl. Tab. 2).

Prior to our analysis, all continuous features and covariates were normalized using z-score normalization. Ordinal covariates such as socio-economic status and mother’s education were scaled between 0 and 1, and gender was treated as a binary variable.

In (Fuzzy) K-Means clustering, all input features are considered equally, and no feature selection is inherently performed. Therefore, it is crucial to carefully choose which features to include, as each will impact the clustering results. To address this, we used principal component analysis (PCA) as a further preprocessing step. PCA helps reduce the dimensionality of the data, retaining only the most significant components based on how much variance is being explained. This is an essential preprocessing step to improve the clustering performance. PCA is suitable for our data type [68, 69], and was applied to all variables. Covariates were also included as control variables, which at the same time allowed us to detect whether any covariate might be influential. Significant principal components were identified using a permutation test (5000 random permutations of each feature across subjects). Components that survived permutation testing were considered significant.

The HCP-EP brain imaging data contained structural magnetic resonance imaging (MRI) data. We used T1-weighted structural images recorded at a 3T SIEMENS MAGNETOM Prisma scanner using an MPRAGE sequence (TR=2400ms, TE=2.22ms, FoV read=256mm, FoV phase=93.8%, flip angle=8 deg, slices per slap=208, slice thickness=0.8mm). In order to receive individual grey matter volume for structural covariance networks generated across all subjects (Suppl. Fig. 2), the structural brain data was preprocessed as described in the supplementary material A.4 and elsewhere [70]. In short, structural covariance network analyses apply an independent component analysis which is based on all subjects’ grey matter maps in order to identify common spatial components. These components are derived from the covariation of grey matter patterns across all participants. We allowed the process to identify 30 components (i.e., structural covariance networks, SCN), as done previously [70–72]. While atlas-based procedures are usually based on brain segmentations derived from healthy brains, the procedure of the present study has the advantage of taking the brain anatomy of both healthy individuals as well as patients into account, thus avoiding a healthy brain bias of other methods. Differences within these group-independent networks identified by the current method should therefore be highly reliable, and the risk of false positive results should be minimized. The structural covariance networks were used for analyses following the clustering.

### 2.4 Correlation and homogeneity

To investigate correlations between all features, we performed a Pearson’s correlation between all subjects’ non-standardized variables. Differences in correlations between the groups were evaluated with a T-test for means of two independent samples. We then tested homogeneity between groups, using Levene’s test for equal variances between groups. Bonferroni corrections were applied.

### 2.5 Fuzzy clustering

Instead of a hard clustering approach that assumes well separated clusters and assigns each data point to only one cluster (e.g., K-Means), we used a soft clustering approach. This approach accounts for fuzzy boundaries between groups, and is thus better suited for overlapping subgroups [73], as would be expected for clinical groups. We, therefore, used Fuzzy K-Means clustering as described in Bezdek et al [74]. As input to our clustering analysis, we used the dimensionality-reduced data which included demographic control variables for all three groups (i.e., controls, affective and non-affective patients). We specified the number of clusters prior to our analysis. We used a priori knowledge about the number of cognitive clusters (i.e., three different cognitive clusters were reported in the literature [54]). As a control analysis, we also used three clusters on the patient data only. This analysis was used to determine influence of control subjects on the clustering, and is presented in the supplements (Suppl. Fig. 4). Although we used a predefined number of clusters, which was based on other studies [54], a data-driven approach supports this view at least partially, showing that three clusters could also be derived from the investigation of inertia (Suppl. Fig. 5). The numbers of clusters should be chosen to achieve low inertia and a low number of clusters. Commonly, the elbow method is employed, which identifies the point after which the improvement in the inertia value levels off. With this method, we identified three clusters the best solution (Suppl. Fig. 5a). However, other partition indices, such as the partition coefficient (Suppl. Fig. 5b) and the partition entropy coefficient (Suppl. Fig. 5c), which are often chosen for fuzzy clustering [75], indicated a two cluster solution as the preferable one as compared to a three or four cluster solution. A larger partition coefficient indicates low overlap between the clusters, meaning better separation, and a larger value of partition entropy coefficient, however, indicates a higher overlap between cluster. As no clear statement can be made from this data-driven point of view, we decided to follow the approach that allowed addressing our research question most appropriately, using a predefined number of three clusters.

Performance of the clustering analysis was determined by the ratio of the subjects of one group (healthy controls, affective psychosis group and non-affective psychosis group) in each cluster. The ratio of subjects in cluster j for group i in {controls, affective and non-affective} is defined as:

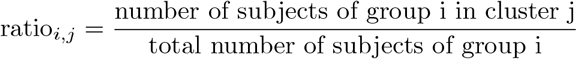

### 2.6 Cluster exploration

After the identification of the three clusters, we explored differences in the patients distributed over those three clusters and controls in cognitive scores, clinical scores, medication and grey matter volume. The control subjects were removed from each of the clusters and combined in one healthy group. Thus, a group comparison across four groups was computed. For comparing groups, we used SciPy’s Kruskal-Wallis test with Dunn’s test for post-hoc analyses, a Chi-square test of independence or ranked analysis of variance (ANOVA) with Bonferroni corrected post-hoc tests. The Kruskal-Wallis test and Dunn’s test for post-hoc analyses are non-parametric, rank-based methods that minimize the impact of outliers by converting data into ranks. This approach reduces the influence of extreme values, as outliers are represented by extreme ranks rather than affecting the analysis based on their raw values. Consequently, both tests offer robust results even in the presence of outliers, ensuring that the outcomes are not disproportionately influenced by these extreme data points. As a control analysis, we performed partial Pearson’s correlations between cognitive scores, clinical scores, and grey matter volume controlled for medication across all patients, with multiple comparison corrections.

### 2.7 Statistical implementation

Preprocessing and data analysis was performed in Python 3.9.7. We used scikit-learn 1.0.2, SciPy 1.7.2 for all analyses and the fuzzy clustering implementation of Dias et al. [76]. Partial correlations were performed using the ppcor 1.1 [77]. For clustering analyses, brain data was corrected for total intracranial volume (TIV), age and sex, using the R stats package, version 4.0.5 R [78].

## 3 Results

### 3.1 Homogeneity of data across groups

Correlations of all cognitive, perceptual and emotional functioning data revealed that correlations were highest within controls compared to affective (T-test: *t*_5838_=16.34, p*<*0.001) and non-affective (T-test: *t*_17058_=26.03, p*<*0.001) patients. For non-affective patients, correlations within subjects were lowest (non-affective vs. affective patients: T-test: *t*_16626_=8.47, p*<*0.001), indicating greatest variability. Confirming these results, highest heterogeneity was found in non-affective patients compared to controls (Levene’s test for equal variance: *F*_1,174_=187.3, p*<*0.001) and affective patients (Levene’s: *F*_1,168_=70.39, p*<*0.001). Controls were the most homogeneous (Levene’s: *F*_1,106_=22.34, p*<*0.001) (Fig. 1a).

**Figure 1:**
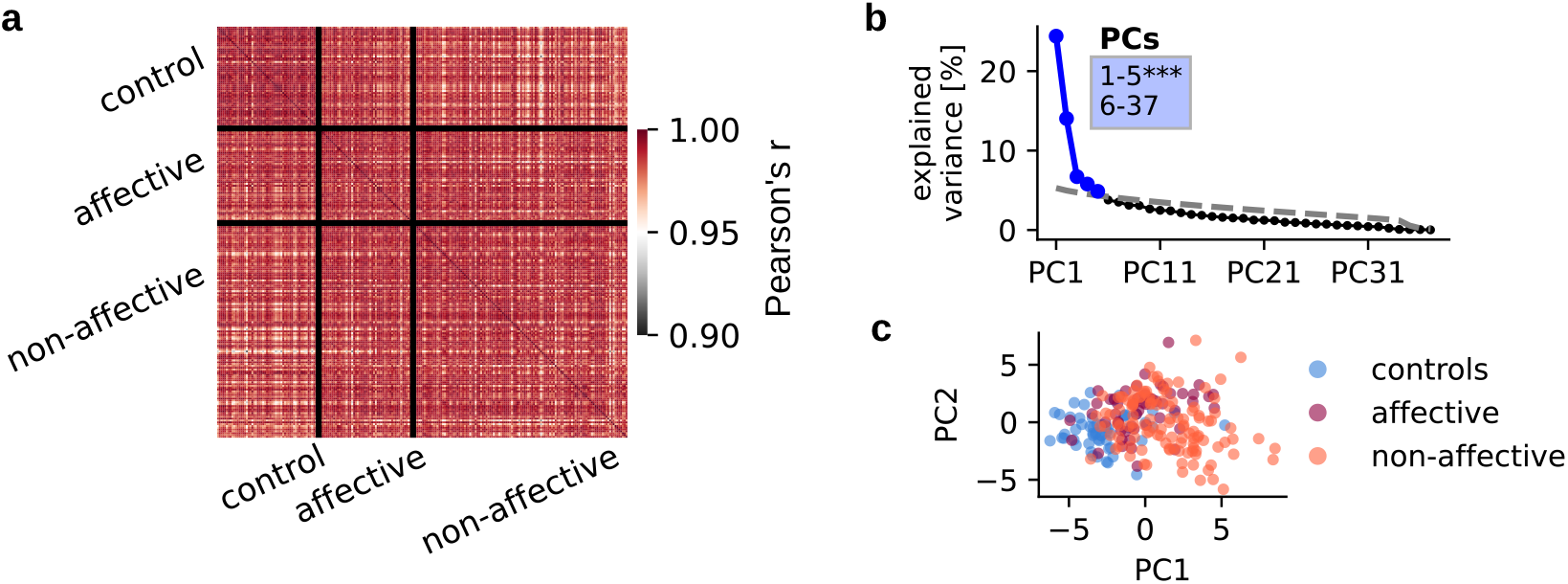
Variable correlations and dimensionality reduction of cognitive data. **(a)** All variables (i.e., cognitive, emotional, perceptual data) correlated across subjects using Pearson correlation. Correlations within groups are displayed in squares on the diagonal, and correlations between groups are displayed in off-diagonal squares. **(b)** Variance explained by each of the principal components (PCs) in % of a PCA performed on all variables and covariates across 226 subjects. The first five PCs (blue) survived permutation testing (p < 0.05, 5000 permutations). Significant components captured 55.8 % of all variance. **(c)** Individual data points represent relevant variables for each subject, displayed on the first two principal components and are colored according to subject group affiliation.

### 3.2 Feature selection using principal component analysis

During the data dimensionality reduction, we identified five significant PCs using all data from all groups, which captured 55.8 % of the total variance (Fig. 1b). Explained variance of PCAs was consistent with other symptom-reduction studies [79]. Fig. 1c illustrates the data reduction using the first two principal components. The two patient groups and the control group are predominantly distributed across the dimension of PC1. However, no clear boundaries between groups could be detected. The features that explained most variance were Fluid Intelligence and Crystallized Intelligence, Total IQ, the Picture Vocabulary Test, Oral reading recognition, Auditory attention, Working memory, WASI - Verbal comprehension, WASI - Matrix reasoning as well as DCCS - Executive functioning (Suppl. Fig. 3). To check the influence of combining patient and control data, the same analysis was performed on only patients data (without controls), which also resulted in five significant components that captured 53.7 % of all variance (p*<*0.05, 5000 permutations) (Suppl. Fig. 4a, b). The top ten features contributing to the explained variance were the same across both analysis - with and without controls. These features were taken for subsequent statistical analyses.

### 3.3 Group representation in the three clusters

We performed clustering on the significant PCs, including controls and both patient groups, with three clusters. Fig. 2a and b present the results. One cluster (cluster 0) contained 86% (48/56) of all controls and 37% (19/52) of all affective subjects. Non-affective subjects were represented only in a small proportion of 17% (20/118). A mixed, second cluster (cluster 1) consisted of mostly patients, with the majority of affective individuals: 54% (28/52) of non-affective and 36% (42/118) of affective patients and only 11% (6/56) controls. Most non-affective subjects were contained in the third cluster (cluster 2) with 47% (56/118) of the non-affective, and only 10% of affective patients (5/52) and 4% (2/56) of controls. (Fig. 2b).

**Figure 2:**
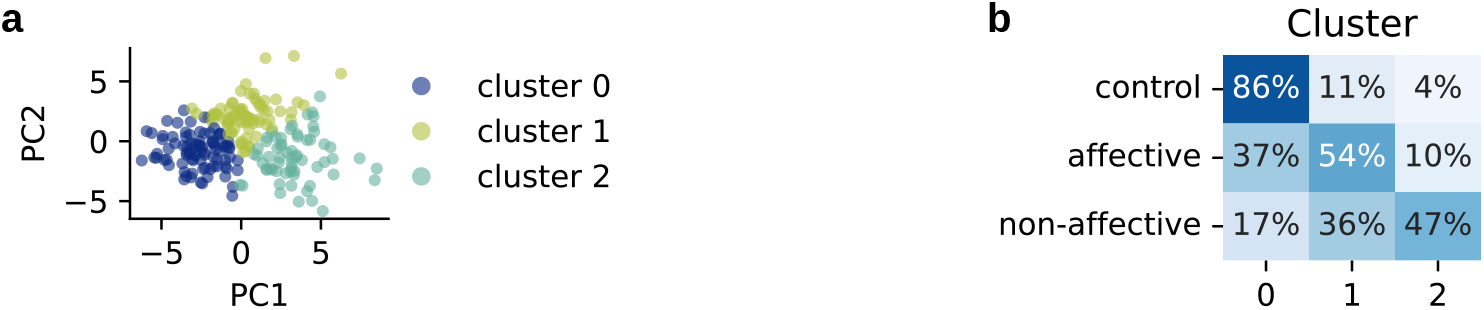
Cluster analysis: group representations and cognitive differences. **(a)** Result of clustering. Cluster affiliation of each subject is displayed on the first two PCs. Colors correspond to cluster 0, 1 or 2. **(b)** Percentage of subjects of each group in each cluster. E.g. 47 % (56/118) of all non-affective subjects are in cluster 2.

### 3.4 Exploring cognition, symptoms, medication and grey matter volume in the clusters

To identify possible cognitive subgroups represented in the clusters, we explored cognitive scores and symptom expression for patients assigned to the clusters (see comprehensive display of statistical differences between all clusters and cognitive items in Suppl. Tab. We compared cognitive scores across patients in respective clusters. Scores of control subjects were used for comparison. Patients in cluster 2 (mainly non-affective patients) showed a significant decrease in all cognitive features compared to patients assigned to other clusters as well as controls. Patients in cluster 1, which contains 28 affective and 42 non-affective patients, showed significant decrease of Fluid Cognition, Auditory attention (%correct) and DCCS - Executive functioning compared to controls. Patients (19 affective and 20 non-affective) in cluster 0 showed most similar scores compared to control subjects, with Auditory attention (%correct) being the only score that was significantly lower compared to controls (Fig. 3c). Group comparisons are presented in Tab. 2.

**Figure 3:**
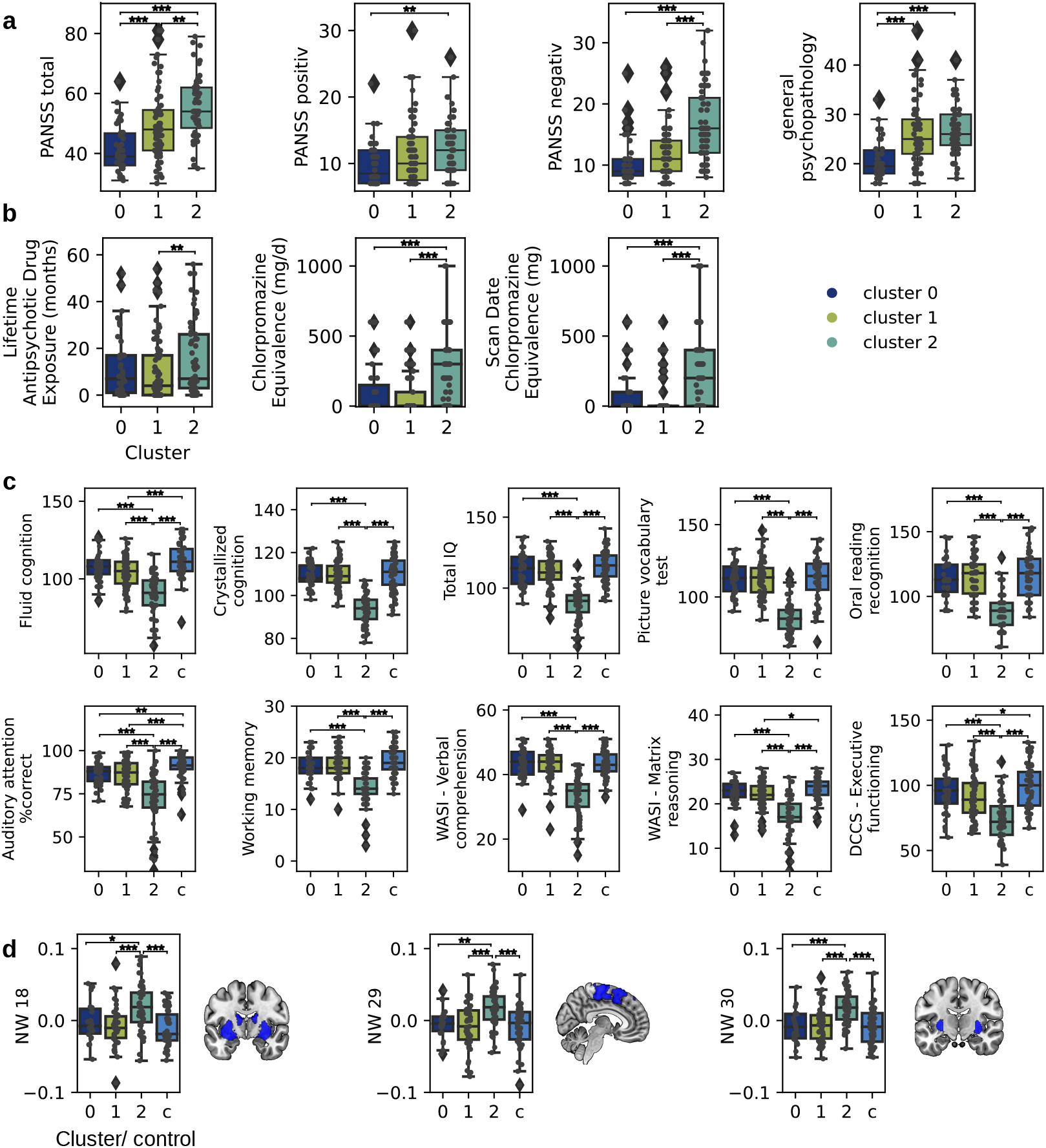
Symptom expression and medication status. **(a)** Clinical scores of affective and non-affective patients within each cluster are displayed in boxplots. Individual boxplots show data minimum, first quartile, median, third quartile, and data maximum. Individual subjects are overlaid as dots. Outliers are indicated outside the data minimum or maximum. **(b)** Medication dosage and status of affective and non-affective patients within each cluster. **(c)** Cognitive scores of affective and non-affective patients within each cluster are displayed and compared to all control subjects (c, light blue). The plots display the ten features contributing most to explained variance of the PCs. **(d)** Differences in three grey matter volume networks between affective and non-affective patients within each cluster are displayed and compared to all control subjects (c, light blue). NW 18 comprises the putamen and the amygdala; NW 29 comprises the paracingulate gyrus, the juxtapositional lobule, the superior parietal lobule, and the precentral gyrus; and the NW 30 comprises the superior frontal gyrus, the frontal pole, the putamen, the postcentral gyrus and the cerebellum crus. The plots display the ten features contributing most to explained variance of the PCs. Significance is indicated as *p* ≤ 0.05 (*), *p* ≤ 0.01 (^**^) and *p* ≤ 0.001 (^***^).

Patients in cluster 2, who showed the strongest cognitive deficits and who were mainly diagnosed with non-affective psychosis, showed a significantly increased PANSS total score compared to patients in both, cluster 0 and 1. They also had a significantly increased PANSS positive, PANSS negative score and general psychopathology compared to patients in cluster 0. Even though patients in cluster 0 and cluster 1 did not show as a strong difference in cognitive scores, patients in cluster 1 had a significantly increased PANSS total score and general psychopathology compared to patients in cluster 0 (Fig. 3a). Group comparisons are presented in Tab. 2.

Further, we explored the medication status of patients within the clusters. We found that patients assigned to cluster 2 had a significantly higher Chlorpromazine equivalence dose in general and at the scanning date compared to patients in both cluster 0 and 1, and a significant increase in lifetime antipsychotic drug exposure compared to patients in cluster 1 (Fig. 3b) and group comparisons are presented in Tab. 2.

We also considered differences in grey matter networks across the patients in the clusters and the controls. The ranked ANOVA revealed a highly significant interaction effect (F(87)=2.564, p=1.79e-13) between the patients in the three clusters and the controls (four groups) and the grey matter volume in each network (30 networks). Bonferroni corrected post-hoc analyses revealed significant differences in network 18 comprising the putamen and the amygdala (F(3,48.64)=9.32, p=0.00027), network 29 consisting of the paracingulate gyrus, the juxtapositional lobule, the superior parietal lobule, and the precentral gyrus (F(3,45.13)=7.8, p=6e-05) and network 30 including the superior frontal gyrus, the frontal pole, the putamen, the postcentral gyrus and the cerebellum crus (F(3,45)=10.95, p=2e-05) (Fig. 3d). Group comparisons are presented in Tab. 2.

Finally, we investigated partial Pearson correlations between the ten cognitive features, the three significant grey matter volume networks, and the clinical scores, controlling for Chlorpromazine equivalent dose within each cluster and across all patients. Across all patients, but not within the clusters, we found several significant, multiple-comparison-corrected associations, see Fig. 4 and Suppl. Tab. 6.

**Figure 4:**
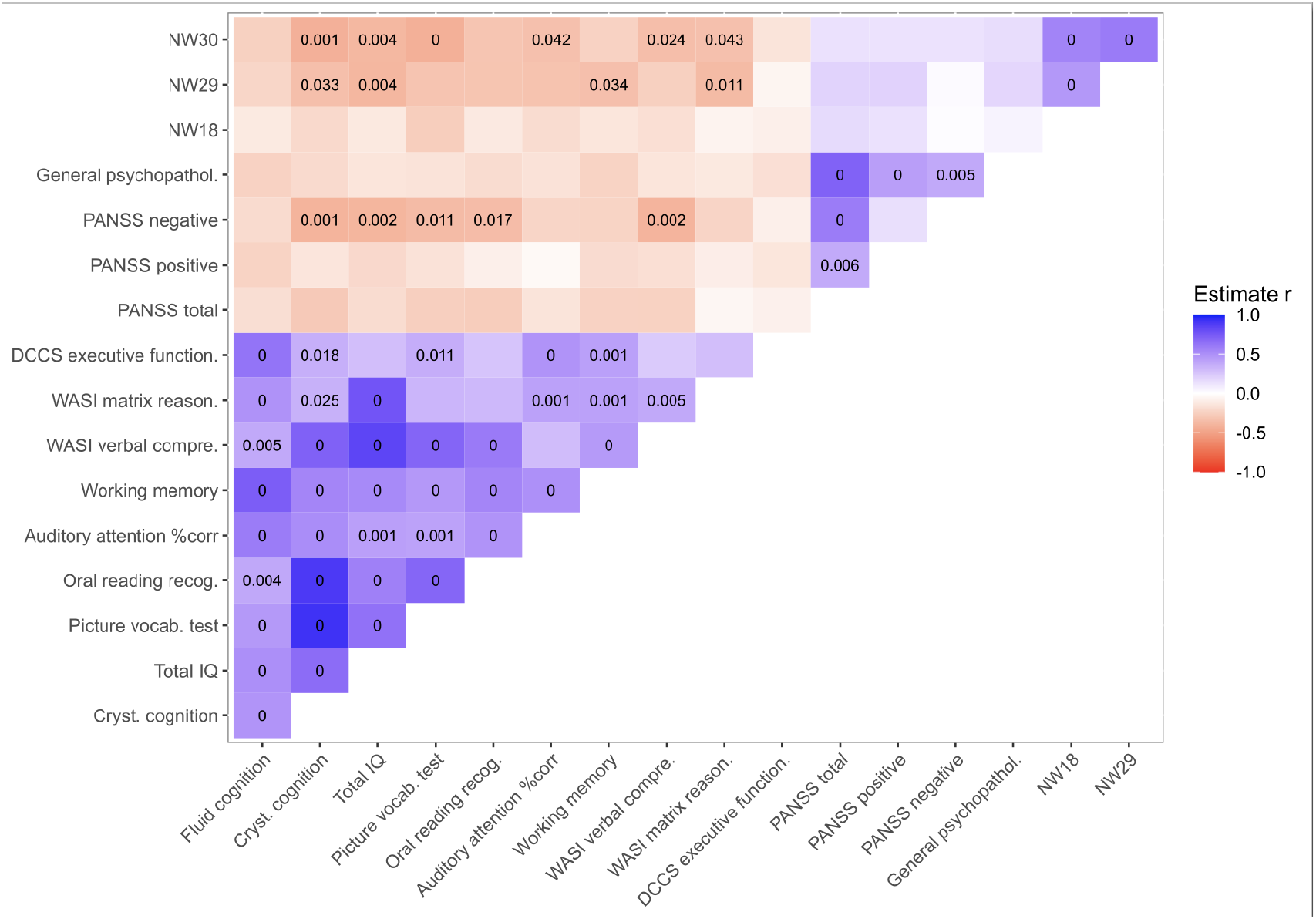
Partial correlations between brain, cognition and clinical scores. **C**orrecting for the equivalent dose of medication, we used Partial Pearson correlations to investigate the interaction between cognitive scores (i.e., Fluid Intelligence and Crystallized Intelligence, Total IQ, the Picture Vocabulary Test, Oral reading recognition, Auditory attention, Working memory, WASI - Verbal comprehension, WASI - Matrix reasoning as well as DCCS - Executive functioning), brain networks (i.e., NW18, NW29, NW30), and clinical scores (i.e., PANSS total, PANSS positive, PANSS negative, General psychopathology score) across all patients independent of cluster.

## 4 Discussion

The aim of this study was, first, to investigate three cognitive subtypes using the HCP Early Psychosis dataset using data-driven clustering on standardized cognitive, perceptual and emotional task and score data, but no clinical data; second, whether the patients in the three clusters differed in symptom expression, cognition, medication and grey matter volume; and, third, depending on the results for the first two aims, we wished to understand if symptoms, alterations in cognition and brain morphometry were associated when controlling for medication within and across the clusters. Using a data driven parameter selection and clustering approach, we were able to show significant differences across our three clusters revealing a cognitively intact cluster, an intermediately affected cluster and a cognitively affected cluster, which seem to confirm subgroups previously described in the literature [36, 52, 80–85]. Importantly, our results extend those findings, showing that patients within those clusters also differ in medication dosage in specific grey matter brain networks and in clinical symptoms. Interestingly, across all patients but not within clusters, we found that decreased grey matter volume in frontal, parietal and subcortical regions was linked to higher cognitive scores including crystallized cognition, verbal comprehension and matrix reasoning, when controlling for medication, and that decreased cognitive scores were linked to increased negative symptoms, when controlled for medication.

Using a three cluster solution on all participants (i.e., controls, affective and non-affective psychosis) allowed the identification of cognitive subtypes, which significantly varied in clinical and cognitive impairment. Patients of cluster 2, consisting of nearly 50% of the nonaffective psychosis individuals, expressed the highest symptom scores across PANSS total, PANSS negative and positive and general psychopathology. They also showed impaired cognition in all domains compared to cluster 0 containing 17% of the non-affective and 37% of the affective individuals, and partially also compared to cluster 1 which consists of the majority of affective individuals and 36% of non-affective individuals. Patients in cluster 0 had the lowest symptom scores and globally spared cognitive abilities, which were similar to those of controls. Cluster 1 was intermediate, with cognitive impairments in several but not all domains and slightly increased symptoms compared to the cognitively spared cluster. This finding confirms results from Lewandowski et al. [52] who found a four-cluster solution to provide the best fit to their data containing three diagnostic patient groups, with one globally impaired cluster for which cognitive deficits were associated with symptom severity and poorer functioning, one cognitively spared cluster and two intermediate clusters [52]. The overall structure of cognitive clusters identified in the present study supports findings discussed in a recent meta-analysis of data-driven identification of cognitive phenotypes in schizophrenia [54]. Green et al. [54] describe that what is characteristic to all cluster solutions is the presence of a cognitively spared, one or multiple intermediate and a deficit subgroup [54]. Importantly, the current study explores three cognitive clusters using standard cognitive, perceptual and emotional assessments, selected for general cognitive screening purposes but not necessarily to detect the largest or most consistent cognitive deficits in early stages of psychosis, suggesting generalisability among these clusters.

In addition to the differences across many cognitive domains between the patients of the clusters, we also found differences with regard to the amount of medication using Chlorpromazin equivalent doses and grey matter volume in frontal, parietal and subcortical brain areas. This emphasizes the complexity of the inter-relationships of cognitive deficits, brain alteration, medication usage and symptom expression, especially when considering that the clustering is based on task and questionnaire data only, and still, differences across all domains - cognition, brain scores, medication and symptoms - have been identified. Several studies indicate an association between higher doses of medication and stronger cognitive deficits [86–88]. In a birth cohort study, for example, Husa et al. [86] showed that a higher lifetime dose of anti-psychotics was associated with lower cognitive performance in schizophrenia patients at the age of 43. Interestingly, a longitudinal study [87] investigating the effect of anti-psychotic treatment discontinuation showed that those individuals who did not remain on their medication after a 3.5 year follow up had improved significantly more than those who stayed on their medication even when controlling for symptom severity and cognitive scores at baseline. General non-adherence of medication use, however, does not have the same positive effect on cognition [89]. Especially, anticholinergic medication has been associated with a high cognitive burden [88], which is supported by our results.

Changes in grey matter volume have been associated with an increased risk for psychosis and disease development [90–92], and provided the basis for good classification in a recent multicohort-study [93], as well as in earlier studies [94, 95], although classification results are inconsistent [96]. Our results show that the cluster with the strongest cognitive deficits has increased grey matter volume in three brain networks spanning fronto-parietal and subcortical areas. Across all participants and when correcting for medication we also found a negative correlation between grey matter volume and cognitive performance in several cognitive tests, including general cognition, verbal cognition and reasoning. Interestingly, grey matter volume alterations, especially reductions have been reported in association with schizophrenia [41, 97–99]. Results however depend on the specific region [100], illness stage and medication [101, 102]. Interactions between grey matter alterations and various cognitive scores have not been studied extensively [103–106]. Most studies investigated cognitive alterations in specific domains (e.g., working memory) and often reported positive correlations [107–109]. Very few studies report negative correlations - increased grey matter volume being linked to reduced cognitive scores. Zhang et al. [110] for example showed, comparable with our results, that the performance in the Stroop Color-Word Test’s Card C was negatively correlated with grey matter volumes of frontal and middle frontal brain areas. In a cohort of at-risk mental state for psychosis individuals, Koutsouleris et al. [105] found positive and negative correlations between grey matter volume and performance in the trail-making test, with negative correlation reported for cerebellar regions, which is comparable to what we report in the current study. We suspect that the structural covariance network analysis, which specifically aims at finding similarity networks between participants, and then analyses grey matter volume differences within these similar networks might contribute to the difference in directionality of the correlations. Our results however clearly demonstrate differences in the cluster containing the most strongly affected patients clinically and cognitively. A study by Wenzel et al. [104] took a different approach. They attempted to classify individuals of different cognitive clusters using grey matter volume data and a support vector machine approach. They achieved a 60.1% classification accuracy, similar to an early study [106], indicating some morphological alterations associated with the cognitive subtypes. Taken together, these differences reported here and in the literature may suggest that grey matter changes are not generally linked to cognitive changes, but rather play a mediating role. This argument would be in the same line of thought as put forward by Palaniyappan [111], stating that grey matter changes in multimodal brain regions which have a supervisory function on sensory, emotional and language processing, may link to symptom expression when occurring with functional impairments. Future studies should therefore aim at the combination of additional structural imaging data, such as structural or white matter connectivity, to complement their analysis and potentially identify underlying neuropathological mechanisms.

Our data furthermore reveals a strong link between negative symptoms and cognitive impairments. Investigating PANSS total, we found a step-wise increase in symptom severity from cluster 0 to cluster 2, with the cognitive deficit cluster (cluster 2) showing highest symptom scores. We found, also, that cluster 0, the cognitively spared cluster, showed significantly lower positive symptoms compared to cluster 1 and 2. Furthermore, the globally impaired cluster (cluster 2) revealed increased negative symptoms compared to both other clusters, and increased global psychopathology compared to the cognitively spared cluster (cluster 0), indicating that the cognitive deficits occur in those subjects with strong negative symptoms and a higher severity of general psychopathology [112]. This is in congruence with Oomen et al. [113] who reported three clusters based on only cognitive data with one severely cognitively impaired cluster, which showed general functioning being significantly lower compared to the patients in the other clusters. Interestingly, patients of the severely impaired cluster also showed lower general functioning scores at a trend at 6- and 12- month follow-up. Similar results were reported by Haining et al. [84]. Tan et al. [85] on the other hand did not find associations between the three cognitive clusters and symptom expression, but found that the cognitively impaired subgroup already showed worse academic performance during childhood, early and late adolescence. These findings confirm the critical relevance of cognitive deficits for early detection and functional prediction [26–28, 84]. This often replicated distribution of a severely impaired cluster, indicating that early interventions based on such cluster analysis would be suitable too.

Finally, as our results confirm that patients with non-affective psychosis show stronger cognitive deficits compared to patients with affective psychosis, it is not surprising that nonaffective patients are more likely to be in the intermediate and impaired subgroups. Still both patient groups are present in all clusters, with the cognitively intact subgroup consisting of 19/52 and 20/118 patients with affective and non-affective psychosis, respectively, the intermediate group with 28/52 affective and 42/118 non-affective psychosis patients, and the cognitively impaired group with 5/52 affective and 56/118 non-affective psychosis patients. Our results indicate that clustering extends classical patient classification and diagnosis solely based on International Statistical Classification of Diseases (ICD)/Diagnostic and Statistical Manual of Mental Disorders (DSM) [114, 115], and may provide an additional characterization of patients which may emphasize additional targets of interventions and treatment, such as cognitive remediation, especially for those individuals in the intermediate and in the deficit cluster. This notion is supported by an increased sensitivity in revealing cluster-specific differences across symptom scores, cognitive scores, grey matter volume and medication, when comparisons between diagnosis groups did not show significant effects (Suppl. Table 2).

**Table 1:** Group demographics and clinical scores of final sample (N=226).

|  | Control | Affective | Non-Affective | Group Comparison<br>KW-chi2/P-ch2,<br>chi2(df), p-value |
| --- | --- | --- | --- | --- |
| N | 56 | 52 | 118 |  |
| Age - mean (std) | 24.55 (4.42) | 23.63 (3.85) | 22.73 (3.38) | 7.25 (2), <0.05 |
| Gender - female % | 33.9 | 57.7 | 30.51 | 11.78 (2), <0.01 |
| PANSS total | na | 45.3 (12.87) | 51.97 (14.54) | 14.71 (1), <0.001 |
| PANSS positiv | na | 10.1 (3.89) | 12.17 (4.59) | 9.09 (1), <0.01 |
| PANSS negative | na | 11.65 (4.66) | 14.14 (5.58) | 9.44 (1), <0.01 |
| General psycho-<br>pathology | na | 23.52 (6.22) | 25.67 (5.69) | 7.01 (1), <0.01 |

**Table 2:** Comparison between patients in all clusters and control subjects for cognitive features that contributed most to variance, as well as medication and PANSS comparison between patients in different clusters. For the group comparison, a Kruskal-Wallis was used with a Dunn’s test for pairwise comparisons (statistics and p-value displayed or ns - non significant).

| Feature | group comparison | cluster 0 - 1 | cluster 0 - 2 | cluster 0 - controls | cluster 1 - 2 | cluster 1 - controls | cluster 2 - controls |
| --- | --- | --- | --- | --- | --- | --- | --- |
| **Cognitive scores** |  |  |  |  |  |  |  |
| Fluid cognition | 87.649, <0.001 | ns | 44.3611, <0.001 | ns | 40.0851, <0.001 | 15.5651, <0.001 | 64.5597, <0.001 |
| Crystallized cognition | 112.1136, <0.001 | ns | 63.2746, <0.001 | ns | 84.5395, ns | ns | 66.9227, <0.001 |
| Total IQ | 108.7698, <0.001 | ns | 57.5106, <0.001 | ns | 74.8416, <0.001 | ns | 73.4878, <0.001 |
| Picture vocabulary test | 101.079, <0.001 | ns | 57.5472, <0.001 | ns | 77.0475, <0.001 | ns | 59.3365, <0.001 |
| Oral reading recognition | 92.0641, <0.001 | ns | 47.5712, <0.001 | ns | 68.125, <0.001 | ns | 59.9688, <0.001 |
| Auditory attention %correct | 74.1838, <0.001 | ns | 31.6126, <0.001 | 11.0381, <0.01 | 33.1923, <0.001 | 12.2936, <0.01 | 57.8224, <0.001 |
| Working memory | 71.9626, <0.001 | ns | 32.4261, <0.001 | ns | 51.8415, <0.001 | ns | 50.0042, <0.001 |
| WASI - Verbal comprehension | 94.1753, <0.001 | ns | 51.1362, <0.001 | ns | 68.2115, <0.001 | ns | 60.9508, <0.001 |
| WASI - Matrix reasoning | 69.0919, <0.001 | ns | 32.4274, <0.001 | ns | 38.7849, <0.001 | 6.9518, <0.05 | 51.9111, <0.001 |
| DCCS - Executive functioning | 54.7055, <0.001 | ns | 27.6009, <0.001 | ns | 27.3806, <0.001 | 5.8383, <0.05 | 42.6152, <0.001 |
| **Medication status** |  |  |  |  |  |  |  |
| Lifetime Antipsychotic Drug Exposure (months) | 7.2749, <0.05 | ns | ns |  | 7.2484, <0.01 |  |  |
| Chlorpromazine Equivalence (mg/d) | 29.132, <0.001 | ns | 12.5926, <0.001 |  | 25.8164, <0.001 |  |  |
| Scan Date Chlorpromazine Equivalence (mg) | 25.9395, <0.001 | ns | 12.1334, <0.001 |  | 22.0764, <0.001 |  |  |
| **Positive and negative symptom score** |  |  |  |  |  |  |  |
| PANSS total | 38.0513, <0.001 | 13.3467, <0.0001 | 35.5926, <0.001 |  | 10.8473, <0.01 |  |  |
| PANSS positive | 8.0705, <0.05 | ns | 8.37, <0.01 |  | ns |  |  |
| PANSS negative | 36.7939, <0.001 | ns | 28.9601, <0.001 |  | 23.4758, <0.001 |  |  |
| general psychopathology | 28.9654, <0.001 | 19.2588, <0.001 | 27.3522, <0.001 |  | ns |  |  |
| **Brain clusters** |  |  |  |  |  |  |  |
| NW 18 | 23.0278, <0.001 | ns | 7.1728, <0.05 | ns | 15.5068, <0.001 | ns | 17.3921, <0.001 |
| NW 29 | 24.305, <0.001 | ns | 11.0358, <0.01 | ns | 15.5068, <0.001 | ns | 17.9004, <0.001 |
| NW 30 | 30.4236, <0.001 | ns | 15.1209, <0.001 | ns | 17.7453, <0.001 | ns | 22.9485, <0.001 |

This study has several limitations: Generally, K-Means [116] is a commonly used clustering algorithm that performed well on our data. Nevertheless, there are some drawbacks of this method: First, the algorithm requires a predefined number of clusters. We did not use a purely data-driven number of clusters. Instead, we predefined three clusters based on previous literature, where a spared, an impaired and an intermediate cluster are regularly detected [54], and the number of three pre-existing diagnosis groups in the data. As a datadriven control analysis for cluster number specification provided non-conclusive evidence for either two or three clusters, we suggest that future research should aim at replicating the current results in a larger cohort and across a more heterogeneous sample of patients, also including chronic schizophrenia patients. Second, K-Means clustering does not work well with non-spherical cluster or clusters with different sizes [117]. We, therefore, also performed fuzzy K-means clustering approach which accounts for fuzzy boundaries between subgroups, and is suited for potentially overlapping subgroups [73]. The age difference between the groups might constitute a third limitation of the study. However, since age was added as a covariate to all analyses, we believe that this aspect did not affect the results to a significant degree. Third, the basis of our clustering approach provided cognitive, perceptual and emotional test scores, which were taken from the standardised NIH toolbox ([63], [64]). Generally, improvement of clustering and identification of cognitive subgroups may be achieved through the selection of specific behavioral tasks and cognitive domains. Moreover, advanced analysis strategies, e.g., computational modelling may improve clustering [16, 17, 118], as it provides the opportunity to identify and mathematically differentiate behavioral parameters, which were found to be reliable and unique across individuals [119].

In conclusion, our results provide evidence for the presence of three cognitive subgroups one cognitively intact, one intermediate and one deficit group - confirming previous findings. This study however extends those results, showing that patients within those three clusters also differ with respect to current medication dosage and grey matter volume in fronto-parietal and subcortical regions. Our results therefore emphasize the complex interrelations between cognition, symptoms, brain structure and medication, drawing attention to the pivotal role of alterations in cognition as a factor for the selection of treatments and interventions.

## Data Availability

Data availability statement: All processed data and code may be accessed via: https://github.com/KatharinaBracher/earlypsychosis_clustering

## Acknowledgement

Research using Human Connectome Project for Early Psychosis (HCP-EP) data reported in this publication was supported by the National Institute of Mental Health of the National Institutes of Health under Award Number U01MH109977. The HCP-EP 1.1 Release data used in this report came from DOI: 10.15154/1522899.

