## Supplements for "Cognitive subgroups of affective and non-affective psychosis show differences in medication and cortico-subcortical brain networks"

Conflict of Interest: None of the authors declares a conflict of interest.

Author contributions: FK, KB, AW conceptualisation; KK preprocessing of structural brain data; FK and KB formal analysis of cognitive data and clustering analysis; FK and KB writing of first draft; FK, KB, AW and KK editing and revising of manuscript.

Acknowledgement: Research using Human Connectome Project for Early Psychosis (HCP-EP) data reported in this publication was supported by the National Institute of Mental Health of the National Institutes of Health under Award Number U01MH109977. The HCP-EP 1.1 Release data used in this report came from DOI: 10.15154/1522899.

Data availability statement: All processed data and code may be accessed via: `https://github.com/KatharinaBracher/earlypsychosis_clustering`

### A Data

#### A.1 Patients

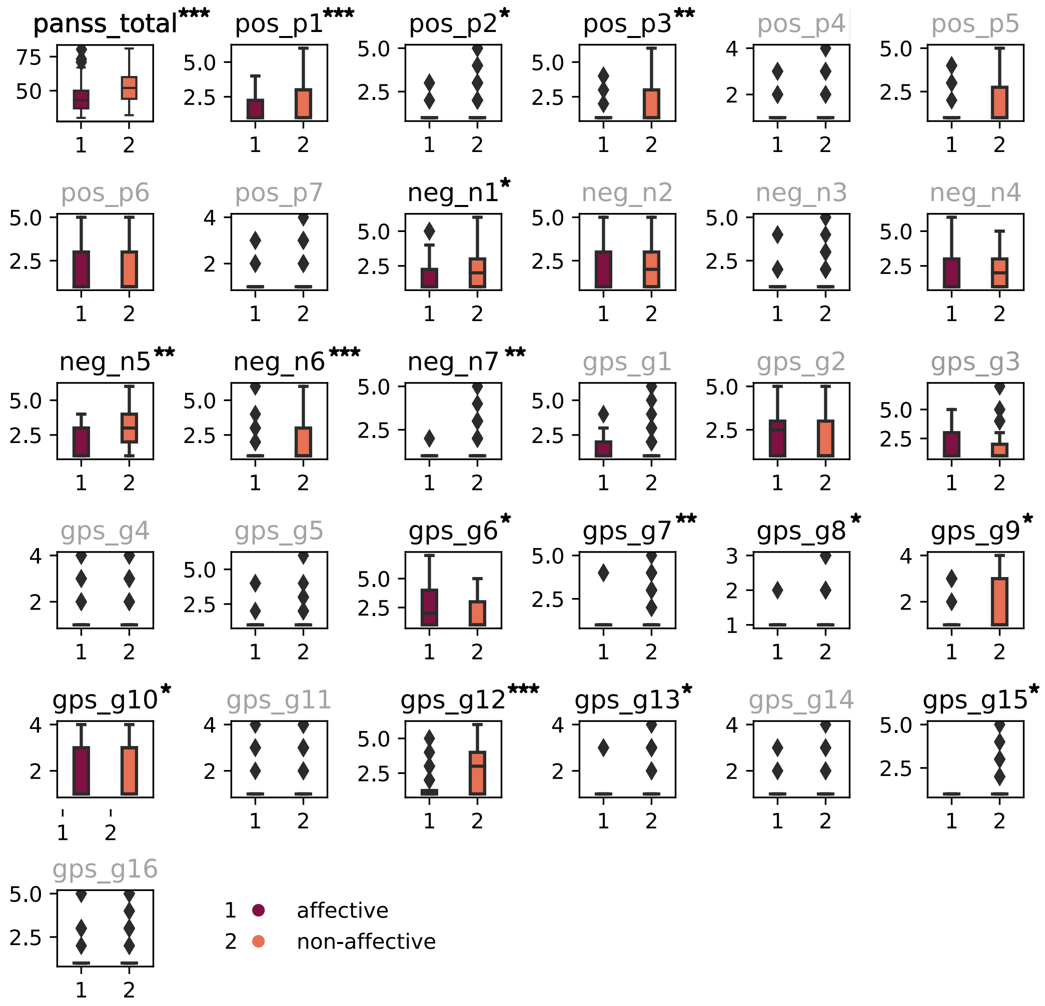


Suppl. Figure 1: **Comparison of patient PANSS items.** Individual boxplots represent the distribution of each PANSS item for affective (1) and non-affective (2) patients, showing data minimum, first quartile, median, third quartile, and data maximum. Outliers are indicated outside the minimum or maximum. Significant differences between affective and non-affective are indicated with stars, where * corresponds to $p < 0.05$, ** to $p < 0.01$ and *** to $p < 0.001$ (using a Kruskal-Wallis H-test).

#### A.2 Cognitive Data

Suppl. Table 1: Features and corresponding description that were considered for our analysis. Of 70 cognitive features, 37 were excluded due to missing entries, which is displayed in the last column where + corresponds to included in analysis and - to excluded of analysis.

| Feature | Description | included |
|---|---|---|
| acpt01_%Hits | Total Q3A-Block : %Hits | + |
| acpt01_RT | Total Q3A-Block : RT | + |
| cgi01_gaf2a | Participant and clinician report-Symptom Scale Score | + |
| cgi01_gaf2b1 | Participant and clinician report-Occupational Functioning Scale Score | + |
| cgi01_gaf2b2 | Participant and clinician report-Occupation | + |
| cgi01_gaf2c | Participant and clinician report-Social Functioning Scale Score | + |
| cogcomp01_nih_fluidcogcomp_unadjusted | Fluid Cognition Composite Score unadjusted | + |
| cogcomp01_nih_crycogcomp_unadjusted | Crystal Cognition Composite Score unadjusted | + |
| dccs01_acc | Accuracy | - |
| dccs01_rt | Reaction Time | - |
| dccs01_nih_dccs_ageadjusted | Age Adjusted scaled score for DCCS subtest | + |
| deldisk01_auc_200 | Area Under the Curve for Delay Amount $200 | + |
| deldisk01_auc_40000 | Area Under the Curve for Delay Amount $40000 | + |
| er4001_er40_c_cr | Correct Responses | - |
| er4001_er40_c_rtcr | Correct Responses Median Response Time (ms) | - |
| er4001_er40_c_ang | Correct Anger Identifications | - |
| er4001_er40_c_fear | Correct Fear Identifications | - |
| er4001_er40_c_hap | Correct Happy Identifications | - |
| er4001_er40_c_noe | Correct Neutral Identifications | - |
| er4001_er40_c_sad | Correct Sad Identifications | - |
| flanker01_acc | Accuracy | - |
| flanker01_rt | Reaction Time | - |
| flanker01_nih_flanker_ageadjusted | Age Adjusted scaled score for Flanker subtest | + |
| lswmt01_nih_tlbx_tscore | T-score | - |
| lswmt01_nih_tlbx_agegencsc | Age-Gender-Corrected T-score | - |
| lswmt01_nih_tlbx_fctsc | Fully-Corrected T-Score | - |
| lswmt01_tbx_ls | List Sort Test total score | + |
| orrt01_read_acss | Reading test Age-Corrected Standard Score | + |
| orrt01_read_fcts | Reading test Fully Corrected T-score | - |
| pcps01_nih_tlbx_fctsc | Fully-Corrected T-Score | - |
| pcps01_nih_patterncomp_ageadjusted | Age Adjusted scaled score for PatternComp subtest | + |
| prang01_anger_ts_nih_toolbox_anger-physical_aggression_ff_age_18+_v2.0 | Anger T score | + |
| prang01_anger_ts_nih_toolbox_anger-hostility_ff_age_18+_v2.0 | Anger T score | + |
| prang01_anger_ts_nih_toolbox_anger-affect_cat_age_18+_v2.0 | Anger T score | + |
| predd01_edd_rs | Emotional Distress Depression raw score | + |
| prsi01_soil_rs | Social Isolation raw score | + |
| psm01_nih_picseq_ageadjusted | Age Adjusted scaled score for PicSeq subtest | + |
| pss01_pss_distress_rs | PSS Distress raw score | + |
| self_effic01_nih_tlbx_rawscore | RawScore | + |
| tlbx_emsup01_nih_tlbx_rawscore_nih_toolbox_instrumental_support_ff_age_18+_v2.0 | RawScore | + |
| tlbx_emsup01_nih_tlbx_rawscore_nih_toolbox_emotional_support_ff_age_18+_v2.0 | RawScore | + |

| Feature | Description | included |
|---|---|---|
| tlbx_friend01_nih_tlbx_rawscore | RawScore | + |
| tlbx_perhost01_nih_tlbx_rawscore | RawScore | + |
| tlbx_rej01_pr_score | Peer Rejection Score | + |
| tlbx_wellbeing01_tlbxpa_ts | Positive affect t-score | + |
| tpvt01_tlbx_readncorr | English Reading Number Correct | - |
| tpvt01_lavoc_screen | Vocabulary Starting theta based on age/education | - |
| tpvt01_tpvt_acss | TPVT age-corrected standard score | + |
| tpvt01_tpvt_fcts | TPVT fully corrected T-score | - |
| wasi201_ss_blockdesigntscoreperf4 | Block Design T - Score Performance - 4 Subtest | - |
| wasi201_ss_vocabularytscoreverbal4 | Vocabulary T - Score Verbal - 4 Subtest | - |
| wasi201_ss_vocabularytscore2 | Vocabulary T - Score - 2 Subtest | - |
| wasi201_ss_matrixreasoningtscoreperf4 | Matrix Reasoning T - Score Performance - 4 Subtest | - |
| wasi201_ss_matrixreasoningtscore2 | Matrix Reasoning T - Score - 2 Subtest | - |
| wasi201_ss_similaritiestscoreverbal4 | Similarities T - Score Verbal - 4 Subtest | - |
| wasi201_sumstscores_verbal4subtest | Verbal - Score - 4 Subtest | - |
| wasi201_sumstscores_perf4subtest | Performance - Score - 4 Subtest | - |
| wasi201_sumstscores_total4subtest | Total - Score - 4 Subtest | - |
| wasi201_sumstscores_total2subtest | Total - Score - 2 Subtest | - |
| wasi201_iqscores_verbsumtscores | Verbal Sum of T - Scores | - |
| wasi201_iqscores_verbiq | Verbal IQ | - |
| wasi201_iqscores_verbpercentile | Verbal Percentile | - |
| wasi201_iqscores_verbconfintervalfrom | Verbal Confidence Interval - From | - |
| wasi201_iqscores_verbconfintervalto | Verbal Confidence Interval - To | - |
| wasi201_iqscores_perfsumtscores | Performance Sum of T - Scores | - |
| wasi201_iqscores_perfiq | Performance IQ | - |
| wasi201_vocab_totalrawscore | Total Raw Score - Vocabulary | + |
| wasi201_matrix_totalrawscore | Total Raw Score - Matrix Reasoning | + |
| wasi201_iqscores_full2iq | Full 2 IQ | + |
| ymrs01_ymrstot | YMRS Total score | - |

Suppl. Table 2: Comparison between patients and control subjects for cognitive features that contributed most to variance, as well as medication and PANSS comparison between diagnosis. For the group comparison, a Kruskal-Wallis was used with a Dunn's test for pairwise comparisons (statistics and p-value displayed or ns - non significant).

| Feature | group comparison | contr. - affect. | contr. - non-aff. | affect. - non-aff. |
|---|---|---|---|---|
| **Cognitive scores** | | | | |
| Fluid cognition | 46.1196, <0.001 | 8.6884, <0.01 | 42.3859, <0.001 | 12.5001, <0.001 |
| Crystallized cognition | 23.732, <0.001 | ns | 20.3015, <0.001 | 8.7481, <0.01 |
| Total IQ | 34.7911, <0.001 | ns | 29.809, <0.001 | 13.3756, <0.001 |
| Picture vocabulary test | 23.1115, <0.001 | ns | 18.5334, <0.001 | 10.6137, <0.01 |
| Oral reading recognition | 17.7379, <0.001 | ns | 16.3271, <0.001 | 4.3483, <0.05 |
| Auditory attention %correct | 43.5029, <0.001 | 10.1902, <0.01 | 40.613, <0.001 | 9.9483, <0.01 |
| Working memory | 30.1556, <0.001 | ns | 22.7316, <0.001 | 16.1795, <0.001 |
| WASI - Verbal comprehension | 19.9352, <0.001 | ns | 16.0612, <0.001 | 9.6666, <0.01 |
| WASI - Matrix reasoning | 34.7254, <0.001 | ns | 30.1583, <0.001 | 12.0238, <0.001 |
| DCCS - Executive functioning | 23.4622, <0.001 | 4.9034, <0.05 | 21.395, <0.001 | 6.2001, <0.05 |
| **Medication status** | | | | |
| Lifetime Antipsychotic Drug Exposure (months) | - | - | - | 24.4946, <0.001 |
| Chlorpromazine Equivalence (mg/d) | - | - | - | 18.7923, <0.001 |
| Scan Date Chlorpromazine Equivalence (mg) | - | - | - | 17.6053, 0.0 |
| **Positiv negative symptom score** | | | | |
| PANSS total | - | - | - | 14.71, <0.001 |
| PANSS positiv | - | - | - | 9.09, <0.01 |
| PANSS negativ | - | - | - | 9.44, <0.01 |
| general psychopathology | - | - | - | 7.01, <0.01 |
| **Brain clusters** | | | | |
| NW 18 | 7.708, <0.05 | ns | 7.4014, <0.01 | ns |
| NW 29 | - | - | - | - |
| NW 30 | 7.9038, <0.05 | ns | 7.7348, <0.01 | ns |

Suppl. Table 3: Features with number of missing individuals. Missing values for continuous features were replaced by the mean according to the group of patients or controls. For categorical data, the mode was used to replace the missing value. *W* stands for 'white', for Socio-Economic Status, 2 for a SES score of 20 to 29 and in the Mother/Father Educational Scale a 4 for High School Graduation or GED and a 6 for Completed 7th through the 9th grades.

| Feature | Missing con. + pat. | Controls mean (std) | Patients mean (std) |
|---|---|---|---|
| **Demographics** | | | |
| Socio-Economic Status | 0+2 | 2 | 2 |
| Mother Educational Scale | 1+6 | 6 | 6 |
| **Auditory continuous performance test** | | | |
| %Hits | 1+3 | 91.0 (6.89) | 81.29 (13.29) |
| Reaction time | 1+3 | 623.19 (72.38) | 669.07 (104.61) |
| **Cognition Composite Scores** | | | |
| Fluid Cognition Composite Score unadjusted | 0+2 | 112.34 (11.22) | 100.07 (13.09) |
| Pattern Comparison Processing Speed - Age adjusted | 0+1 | 104.32 (20.44) | 91.54 (21.08) |
| **Delay Discounting Task** | | | |
| Area Under the Curve Delay Amount $200 | 0+1 | 0.27 (0.26) | 0.25 (0.26) |
| Area Under the Curve for Delay Amount $40000 | 0+1 | 0.55 (0.28) | 0.43 (0.32) |
| **PROMIS Anger** | | | |
| Physical Aggression Anger T score | 0+3 | 49.25 (9.15) | 55.65 (11.17) |
| Hostility Anger T score | 0+3 | 52.14 (9.45) | 57.68 (10.21) |
| Affect Anger T score | 0+3 | 45.8 (10.62) | 50.94 (14.2) |
| **NIH Toolbox Sadness CAT Age 18+ v2.0** | | | |
| Emotional Distress Depression raw score | 0+3 | 9.34 (2.55) | 11.86 (6.74) |
| **NIH Toolbox Sadness CAT Age 18+ v2.0** | | | |
| Age Adjusted scaled score for PicSeq subtest | 0+1 | 106.71 (14.16) | 92.91 (15.27) |
| **NIH Toolbox Sadness CAT Age 18+ v2.0** | | | |
| Instrumental Support Raw Score | 0+3 | 32.29 (6.27) | 29.46 (8.36) |
| Emotional Support Raw Score | 0+3 | 34.25 (7.12) | 30.98 (7.07) |
| **NIH Toolbox Emotion Domain - Psychological Well-Being** | | | |
| Positive affect t-score | 0+3 | 49.45 (6.9) | 43.55 (8.82) |

Suppl. Table 4: Number of missing features per subject

| Subject ID | Number of missing features |
|---|---|
| 1057 | 1 |
| 1063 | 4 |
| 1068 | 2 |
| 1099 | 1 |
| 1118 | 2 |
| 2005 | 7 |
| 2007 | 7 |
| 2012 | 2 |
| 2023 | 7 |
| 3031 | 2 |
| 4053 | 1 |
| 4059 | 1 |
| 4063 | 1 |
| 4065 | 2 |
| 4092 | 2 |
| 1015 | 2 |

#### A.3 Brain Data

**NW1**:

1: 6932, 14, -66, -48, cerebellum VIIIa

2: 6590, -20, -69, -60, cerebellum VIIIa, cerebellum crus

**NW2**:

1: 6763, 8, -48, -28, cerebellum VI

2: 5574, -33, -51, -39, cerebellum VI

**NW3**:

1: 7340, -18, -70, -28, cerebellum VI, cerebellum crus I

**NW4**:

1: 11747, 2, -57, -27, cerebellum I-V

2: 1168, -16, -40, -58, cerebellum VIIIb

3: 134, 21, -42, -56, cerebellum VIIIb

**NW5**:

1: 10338, 6, -74, -38, cerebellum VIIb, cerebellum crus

**NW6**:

1: 5836, -18, -2, -33, parahippocampal gyrus, temporal pole

2: 3948, 34, -6, -51, parahippocampal gyrus, temporal pole

3: 329, -21, -87, -46, cerebellum crus II

**NW7**:

1: 10263, -2, -56, -46, r. cerebellum IX, l. cerebellum IX

2: 489, -27, -56, -36, cerebellum VI, cerebellum V

**NW8**:

1: 6947, -42, -60, -56, cerebellum VIIb, cerebellum crus

2: 5665, 40, -58, -51, cerebellum VIIb, cerebellum crus

3: 352, -14, -6, 10, thalamus

**NW9**:

1: 6039, -40, -4, -38, fusiform, middle temporal gyrus, inferior temporal gyrus

2: 5371, 48, -10, -46, middle temporal gyrus, inferior temporal gyrus

**NW10**:

1: 6981, 3, -78, 8, intracalcarine cortex, lingual gyrus

2: 177, 32, -82, 26 lateral occipital cortex

**NW11**:

1: 12704, 0, 20, 27, cingulate gyrus

2: 176, -33, 33, 33, middle frontal gyrus

3: 109, 34, 27, 30, middle frontal gyrus

4: 100, -8, -4, 15, thalamus

**NW12**:

1: 7408, 2, -64, 18, precuneus, supracalcarine cortex

**NW13**:

1: 5803, -8, -66, 22, precuneus

2: 1015, -28, -62, -33, cerebellum VI, cerebellum crus I

3: 689, 33, -26, 51, postcentral gyrus, precentral gyrus

4: 192, 33, 6, -4, insula

5: 148, 32, -6, -44, fusiform gyrus

6: 112, 36, -36, 39, supramarginal gyrus, superior parietal cortex

**NW14**:

1: 3720, 6, -57, 4, lingual gyrus, precuneus

2: 2335, -14, -48,-9, lingual gyrus, posterior cingulate

**NW15**:

1: 3778, 14, -9, -21, hippocampus, parahippocampal gyrus

2: 3215, -24, -4, -36, hippocampus, parahippocampal gyrus

Soll ich den migratino plot 3: 767, 16, -66, 20, n. accumbens

4: 197, 16, -66, 20, cuneal cortex, supracalcarine cortex

**NW16**:

1: 25138, -4, 48, 3, paracingulate gyrus, cingulate gyrus, middle frontal gyrus, frontal pole

**NW17**:

1: 9529, 0, 28, -14, subcallosal cortex, medial frontal cortex

**NW18**:

1: 8174, -3, 4, -2, putamen, amygdala

2: 7660, -3, 4, -2, putamen, amygdala

**NW19**:

1: 3418, 12, -96, 3, occipital pole
2: 2974, -14, -99, -14, occipital pole
3: 510, 39, -64, -48, cerebellum crus
**NW20**:
1: 3082, -20, 22, 2, caudate, insula
2: 2833, 51, 30, -14, orbitofrontal cortex, insula
**NW21**:
1: 3223, 28, -18, -32, parahippocampal gyrus, fusiform gyrus
2: 2397, -32, -10, -48, fusiform gyrus
3: 339, 48, -52, -18, inferior temporal gyrus
**NW22**:
1: 5026, -12, 10, -18, temporal pole, orbitofrontal cortex
2: 3925, 30, 10, -30, temporal pole, orbitofrontal cortex
3: 2263, 38, -12, -14, insula
4: 1655, -66, -20, -4, superior temporal gyrus, planum temporale
5: 631, -30, -52, -56, cerebellum VIIIa
**NW23**:
1: 2188, 33, -33, 40, postcentral gyrus
2: 1265, -64, -22, 26, postcentral gyrus
3: 172, -50, -27, 10, Heschl gyrus, planum temporale
4: 142, -54, -10, 2, Heschl gyrus, planum temporale
5: 141, -20, -69, -18, cerebellum VI
**NW24**:
1: 6802, -34, -48, -10, inferior temporal gyrus, middle temporal gyrus, fusiform gyrus
2: 4781, 57, -46, -24, inferior temporal gyrus, middle temporal gyrus
3: 406, -38, -58, -57, cerebellum VIIb, cerebellum VIIIa
4: 144, 44, 27, -9, orbitofrontal cortex
5: 32, 26, -24, orbitofrontal cortex, temporal pole
**NW25**:
1: 992, 21, -75, -18, cerebellum VI
2: 316, 22, -84, -38, cerebellum crus
3: 213, 56, -2, -32, middle temporal gyrus
4: 133, 33, -27, -30, fusiform gyrus
5: 126, 40, 10, 38, temporal pole
**NW26**:
1: 10618, -2, -20, 4, thalamus
2: 476, -21, -87, -46, cerebellum crus II
3: 462, -30, -15, 64, precentral gyrus

4: 115, -33, -96, -6, occipital pole

**NW27**:

1: 5041, -4, -48, 24, cingulate gyrus

2: 764, -32, -62, -60, cerebellum VIIIa

3: 188, 12, -74, 22, precuneus

**NW28**:

1: 4057, 51, -15, -18, middle temporal gyrus

2: 1945, -51, -16, -20, middle temporal gyrus

3: 407, 20, -80, -32, cerebellum crus

4: 292, -46, 8, -3, frontal operculum

**NW29**:

1: 8736, -3, 8, 58, paracingulate gyrus, juxtapositional lobule

2: 177, -27, -51, 60, superior parietal lobule

3: 125, 26, -14, 56, precentral gyrus

**NW30**:

1: 2731, -22, 32, 32, superior frontal gyrus, frontal pole

2: 2456, 26, 57, -8, frontal pole

3: 910, 32, -16, -10, putamen

4: 761, -30, -18, -6, putamen

5: 611, 22, -75, -30, cerebellum crus

6: 177, -16, 4, 58, superior frontal gyrus

7: 122, -38, -27, 42, postcentral gyrus

#### A.4 Preprocessing Brain Data

The structural images of all subjects were segmented into grey matter, white matter, and CSF using Statistical Parametric Mapping, running on MATLAB version 2018b. We used Diffeomorphic Anatomical Registration through Exponentiated Lie Algebra toolbox (DARTEL) [1] to process grey matter images. This procedure creates a sample-specific template representative of all subjects by iteratively aligning all images. Then, the template underwent non-linear registration with modulation for linear and non-linear deformations to the MNI-ICBM152 template. Subsequently, we registered each participant's grey matter map to the group template and smoothed with an 8 $mm^3$ isotropic Gaussian kernel.

Following the preprocessing, we computed an independent component analysis. First, all individually preprocessed grey matter maps were concatenated, creating a 4D file. An absolute grey matter threshold of 0.1 was applied to all images, ensuring that only grey matter voxels were used for the ICA. ICA was performed using the Multivariate Exploratory Linear Optimized Decomposition into Independent Components (MELODIC) method as implemented in the FSL analysis package jenkinson2012fsl version 6.0. Data-driven population-based networks of grey matter covariance were derived, performing an ICA on all subjects (n=180). Therefore, this process identifies common spatial components based on the co-variation of grey matter patterns across all subjects. We allowed the process to identify 30 components (i.e., structural covariance networks, SCN), as done previously [2–4]. The results were thresholded at z = 3.5 and binarized [5, 6] to eliminate spurious results. Finally, for each participant, grey matter volume was extracted from each of the 30 morphometric networks.

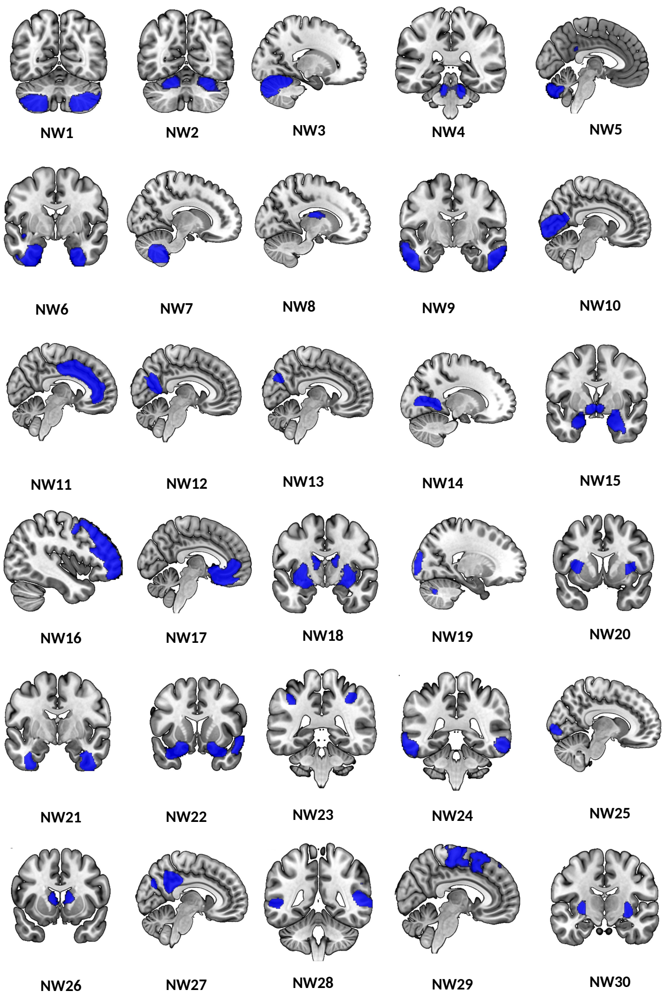


Suppl. Figure 2: **Morphometric grey matter networks.** In this figure, 30 anatomically derived morphometric networks from the ICA thresholded at Z>3.5 are displayed. Each network identifies structural grey matter covariance across all subjects.

### B Results

#### B.1 Feature Selection

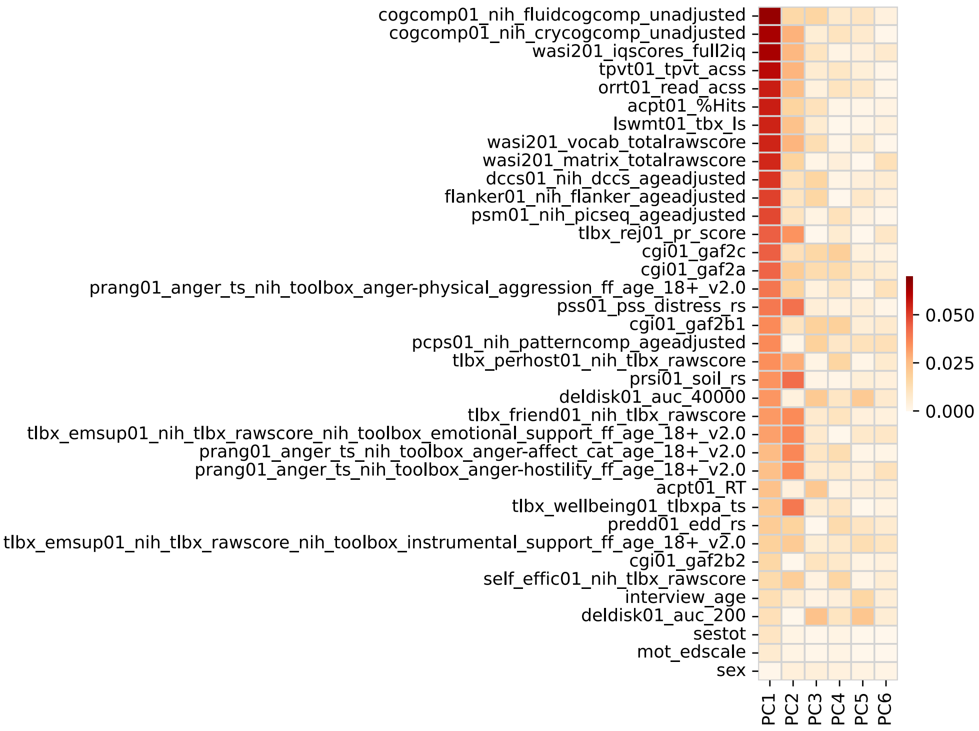


Suppl. Figure 3: **PC loadings of cognitive, perceptual and emotional data.** PCA was performed across 226 subjects on 33 features plus 4 control variables features. Displayed are PC loadings, each normalized with variance explained by the PC, making them comparable across components.

#### B.2 Analysis Excluding Control Subjects

Excluding controls from the clustering resulted in an overall similar distribution of subjects across the clusters (Suppl. Fig. 4), with 151/182 remaining in the same clusters. Interestingly, a part of the non-affective subjects assigned to cluster 1 were instead assigned to cluster 0 when clustering without control, possibly indicating a lower threshold for cognitive integrity. This means that individuals with more subtle cognitive deficits would also be clustered in the cognitively intact cluster. This emphasises the benefit of including controls into the clustering, as it allows true identification of those patients with intact cognition. Cluster 2 remained overall very stable (Suppl. Fig. 4c-e).

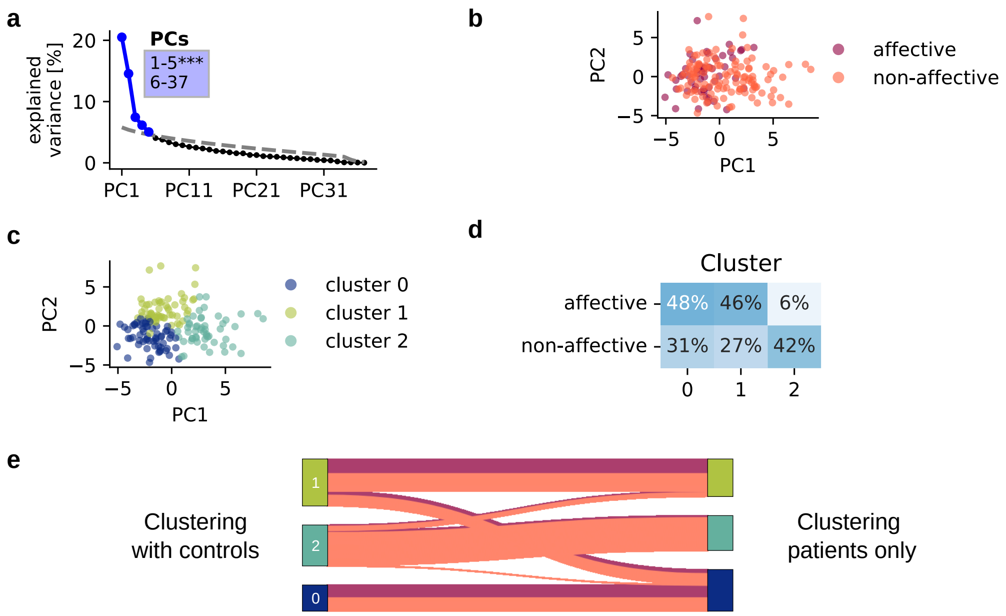


Suppl. Figure 4: **Dimensionality reduction of cognitive and brain data with patients only.** **(a)** Variance explained by each of the PCs in % of a PCA performed on all cognitive features and control variables across 170 patients. The first five PCs (blue) survived permutation testing ($p<0.05$, 5000 permutations). Significant components captured 53.7 % of all variance. **(b)** Cognitive data is displayed on the first two principal components and colored according to patient group affiliation. **(c)** Result of clustering that was performed on cognitive data on patients only with three clusters. Cluster affiliation of each subject is displayed on the first two PCs. Colors correspond to cluster 0, 1 or 2. **(d)** Percentage of subjects of a group in each cluster. E.g. 42 % of all non-affective subjects are in cluster 2. **(e)** Transition graph shows the transitions of individuals between clusters based on cognitive data including control (left) and cognitive of patients only (right). Subjects are divided into assigned clusters, respectively. A line connects each individual between both clusterings. The line is colour coded according to patient group (see legend in b). The graph shows the stability of the clustering.

#### B.3 Cluster Validation Analysis

We validated the chosen three cluster approach using a data-driven approach. Here, the numbers of clusters should be chosen to achieve low inertia and a low number of clusters. Commonly, the elbow method is employed, which identifies the point after which the improvement in the inertia value levels off. With this method, three clusters are indeed identified as the best solution (Suppl. Fig. 5a). However, other partition indices such as the partition coefficient(Suppl. Fig. 5b) and the partition entropy coefficient (Suppl. Fig. 5c), which are often chosen for fuzzy clustering [7], indicated a two cluster solution as the preferable one as compared to a three or four cluster solution. A larger partition coefficient indicates low overlap between the clusters, hence better separation, and a larger value of partition entropy coefficient however indicates a higher overlap between cluster. As no clear statement can be made from this data-driven point of view, we felt confident to follow the approach that allowed addressing our research question most appropriately, using a predefined number of three clusters.

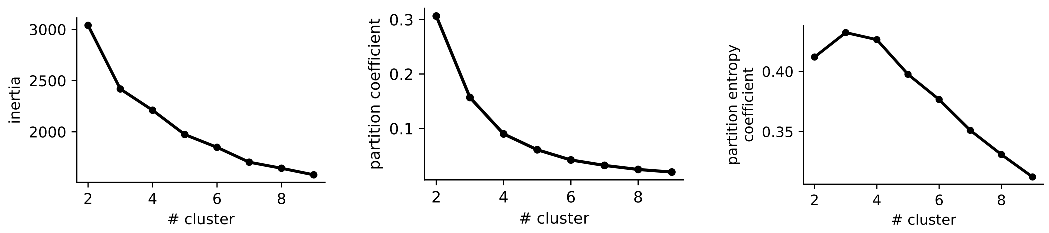


(a) (b) (c)

Suppl. Figure 5: Clustering on the five significant PCs, including controls and both patient groups, with different numbers of clusters. (a) Inertia is the sum of squared distances of samples to their closest cluster centre. The number of clusters should be chosen to achieve low inertia and a low number of clusters. (b) Partition coefficient measures the quality of a fuzzy partition. A large value means well separated clusters. (c) The partition entropy coefficient indicates fuzziness. A large value indicates high overlap between clusters.

#### B.4 Cluster Exploration

Suppl. Table 5: Comparison between patients in all clusters and control subjects for cognitive features. For the group comparison, a Kruskal-Wallis was used with a Dunn's test for pairwise comparisons (statistics and p-value displayed or ns - non-significant).

| Feature | group comparison | cluster 0 - 1 | cluster 0 - 2 | cluster 0 - controls | cluster 1 - 2 | cluster 1 - controls | cluster 2 - controls |
|---|---|---|---|---|---|---|---|
| cgi01_gaf2a | 90.9585, <0.001 | 22.1896, <0.001 | 16.8475, <0.001 | 16.4083, <0.01 | ns | 64.5288, <0.001 | 57.9353, <0.001 |
| cgi01_gaf2b1 | 78.1465, <0.001 | 14.2375, <0.001 | 13.3187, <0.001 | 17.5608, <0.001 | ns | 53.5162, <0.001 | 55.026, <0.001 |
| cgi01_gaf2b2 | ns | - | - | - | - | - | - |
| cgi01_gaf2c | 90.8192, <0.001 | 27.045, <0.001 | 32.1442, <0.001 | ns | ns | 55.1229, <0.001 | 57.138, <0.001 |
| cogcomp01_nih_fluidcogcomp_unadjusted | 87.649, <0.001 | ns | 44.3611, <0.001 | ns | 40.0851, <0.001 | 15.5651, <0.001 | 64.5597, <0.001 |
| cogcomp01_nih_crycogcomp_unadjusted | 112.1136, <0.001 | ns | 63.2746, <0.001 | ns | 84.5395, <0.001 | ns | 66.9227, <0.001 |
| dccs01_nih_dccs_ageadjusted | 54.7055, <0.001 | ns | 27.6009, <0.001 | ns | 27.3806, <0.001 | 5.8383, <0.05 | 42.6152, <0.001 |
| deldisk01_auc_200 | 14.7643, <0.01 | ns | 8.8539, <0.001 | ns | 8.2377, <0.01 | ns | 10.2426, <0.01 |
| deldisk01_auc_40000 | 30.8906, <0.001 | ns | 17.4767, <0.001 | ns | 14.8141, <0.001 | ns | 24.9015, <0.001 |
| flanker01_nih_flanker_ageadjusted | 64.5138, <0.001 | ns | 29.5959, <0.001 | ns | 26.6117, <0.001 | 14.8705, <0.001 | 49.07, <0.001 |
| lswmt01_tbx_ls | 71.9626, <0.001 | ns | 32.4261, <0.001 | ns | 51.8415, <0.001 | ns | 50.0042, <0.001 |
| orrt01_read_acss | 92.0641, <0.001 | ns | 47.5712, <0.001 | ns | 68.125, <0.001 | ns | 59.9688, <0.001 |
| pcps01_nih_patterncomp_ageadjusted | 21.3653, <0.001 | ns | 6.1977, <0.05 | ns | ns | 10.6352, <0.01 | 18.9038, <0.001 |
| prang01_anger_ts_nih_toolbox_anger-physical_aggression_ff_age_18+_v2.0 | 36.5683, <0.001 | 10.8235, <0.01 | 21.9381, <0.001 | ns | ns | 12.3361, <0.001 | 24.0889, <0.001 |
| prang01_anger_ts_nih_toolbox_anger-hostility_ff_age_18+_v2.0 | 32.8018, <0.001 | 18.9004, <0.001 | ns | ns | 6.3975, <0.01 | 27.7908, <0.001 | 5.5897, <0.05 |
| prang01_anger_ts_nih_toolbox_anger-affect_cat_age_18+_v2.0 | 37.5825, <0.001 | 30.3131, <0.001 | 3.7047, <0.05 | ns | 10.285, <0.001 | 25.3552, <0.001 | ns |
| predd01_edd_rs | 33.6906, <0.001 | 21.8731, <0.001 | 11.644, <0.01 | ns | ns | 20.7526, <0.001 | 8.3949, <0.01 |
| prsi01_soil_rs | 77.9705, <0.001 | 40.4617, <0.001 | ns | ns | 25.999, <0.001 | 61.7518, <0.001 | 11.7293, <0.001 |
| psm01_nih_picseq_ageadjusted | 61.2061, <0.001 | ns | 20.3563, <0.001 | 5.7574, <0.05 | 20.8384, <0.001 | 18.307, <0.001 | 51.7783, <0.001 |
| pss01_pss_distress_rs | 69.9825, <0.001 | 34.9998, <0.001 | 15.2583, <0.001 | ns | 8.5161, <0.05 | 49.2708, <0.001 | 25.7544, <0.001 |
| self_effic01_nih_tlbx_rawscore | 21.5996, <0.001 | 9.3138, <0.01 | 3.8954, <0.05 | ns | ns | 18.1286, <0.001 | <0.01 |
| tlbx_emsup01_nih_tlbx_rawscore_nih_toolbox_instrumental_support_ff_age_18+_v2.0 | 8.8109, <0.05 | 4.7407, <0.05 | ns | ns | ns | 7.57, <0.01 | ns |
| tlbx_emsup01_nih_tlbx_rawscore_nih_toolbox_emotional_support_ff_age_18+_v2.0 | 53.608, <0.001 | 47.2719, <0.001 | 8.0771, <0.01 | ns | 12.9624, <0.001 | 30.7682, <0.001 | 5.0836, <0.05 |
| tlbx_friend01_nih_tlbx_rawscore | 68.0718, <0.001 | 38.4971, <0.001 | 9.7359, <0.01 | ns | 9.4773, <0.01 | 54.3892, <0.001 | 17.1694, <0.001 |
| tlbx_perhost01_nih_tlbx_rawscore | 32.9081, <0.001 | 28.8124, <0.001 | 6.5496, <0.01 | ns | 3.7249, <0.05 | 20.8062, <0.001 | ns |
| tlbx_rej01_pr_score | 60.2871, <0.001 | 44.1868, <0.001 | 16.6701, <0.001 | ns | 4.5307, <0.05 | 38.9551, <0.001 | 12.9837, <0.001 |
| tlbx_wellbeing01_tlbxpa_ts | 65.6581, <0.001 | 36.564, <0.001 | ns | ns | 25.2368, <0.001 | 50.8968, <0.001 | 6.581, <0.05 |
| tpvt01_tpvt_acss | 101.079, <0.001 | ns | 57.5472, <0.001 | ns | 77.0475, <0.001 | ns | 59.3365, <0.001 |
| wasi201_vocab_totalrawscore | 94.1753, <0.001 | ns | 51.1362, <0.001 | ns | 68.2115, <0.001 | ns | 60.9508, <0.001 |
| wasi201_matrix_totalrawscore | 69.0919, <0.001 | ns | 32.4274, <0.001 | ns | 38.7849, <0.001 | 6.9518, <0.05 | 51.9111, <0.001 |
| wasi201_iqscores_full2iq | 108.7698, <0.001 | ns | 57.5106, <0.001 | ns | 74.8416, <0.001 | ns | 73.4878, <0.001 |
| acpt01_%Hits | 74.1838, <0.001 | ns | 31.6126, <0.001 | 11.0381, <0.01 | 33.1923, <0.001 | 12.2936, <0.001 | 57.8224, <0.001 |
| acpt01_RT | 11.039, <0.05 | ns | ns | ns | ns | 4.5509, <0.05 | 10.3635, <0.001 |

Suppl. Table 6: Partial correlations between ten cognitive scores, three brain networks, and four clinical scores, across all patients corrected for medication; p-values are corrected for multiple comparisons.

| V1 | V2 | estimate | statistic | p.value | adj_p.value |
|---|---|---|---|---|---|
| Fluid cognition | Cryst. cognition | 0.48 | 5.66 | 0.00 | 0.00 |
| Fluid cognition | Total IQ | 0.50 | 6.03 | 0.00 | 0.00 |
| Fluid cognition | Picture vocab. test | 0.46 | 5.37 | 0.00 | 0.00 |
| Fluid cognition | Oral reading recog. | 0.39 | 4.38 | 0.00 | 0.00 |
| Fluid cognition | Auditory attention %corr | 0.59 | 7.53 | 0.00 | 0.00 |
| Fluid cognition | Working memory | 0.73 | 11.17 | 0.00 | 0.00 |
| Fluid cognition | WASI verbal compre. | 0.38 | 4.31 | 0.00 | 0.00 |
| Fluid cognition | WASI matrix reason. | 0.49 | 5.82 | 0.00 | 0.00 |
| Fluid cognition | DCCS executive function. | 0.63 | 8.38 | 0.00 | 0.00 |
| Fluid cognition | PANSS total | -0.18 | -1.87 | 0.06 | 1.00 |
| Fluid cognition | PANSS positive | -0.24 | -2.55 | 0.01 | 1.00 |
| Fluid cognition | PANSS negative | -0.20 | -2.07 | 0.04 | 1.00 |
| Fluid cognition | General psychopathol. | -0.24 | -2.56 | 0.01 | 1.00 |
| Fluid cognition | NW18 | -0.12 | -1.23 | 0.22 | 1.00 |
| Fluid cognition | NW29 | -0.22 | -2.34 | 0.02 | 1.00 |
| Fluid cognition | NW30 | -0.25 | -2.72 | 0.01 | 1.00 |
| Cryst. cognition | Total IQ | 0.66 | 9.19 | 0.00 | 0.00 |
| Cryst. cognition | Picture vocab. test | 0.92 | 25.21 | 0.00 | 0.00 |
| Cryst. cognition | Oral reading recog. | 0.89 | 20.83 | 0.00 | 0.00 |
| Cryst. cognition | Auditory attention %corr | 0.51 | 6.16 | 0.00 | 0.00 |
| Cryst. cognition | Working memory | 0.54 | 6.72 | 0.00 | 0.00 |
| Cryst. cognition | WASI verbal compre. | 0.71 | 10.61 | 0.00 | 0.00 |
| Cryst. cognition | WASI matrix reason. | 0.35 | 3.88 | 0.00 | 0.02 |
| Cryst. cognition | DCCS executive function. | 0.36 | 3.96 | 0.00 | 0.02 |
| Cryst. cognition | PANSS total | -0.29 | -3.16 | 0.00 | 0.28 |
| Cryst. cognition | PANSS positive | -0.14 | -1.46 | 0.15 | 1.00 |
| Cryst. cognition | PANSS negative | -0.41 | -4.63 | 0.00 | 0.00 |
| Cryst. cognition | General psychopathol. | -0.20 | -2.09 | 0.04 | 1.00 |
| Cryst. cognition | NW18 | -0.20 | -2.16 | 0.03 | 1.00 |
| Cryst. cognition | NW29 | -0.34 | -3.80 | 0.00 | 0.03 |
| Cryst. cognition | NW30 | -0.42 | -4.87 | 0.00 | 0.00 |
| Total IQ | Picture vocab. test | 0.64 | 8.65 | 0.00 | 0.00 |
| Total IQ | Oral reading recog. | 0.57 | 7.19 | 0.00 | 0.00 |
| Total IQ | Auditory attention %corr | 0.43 | 4.88 | 0.00 | 0.00 |
| Total IQ | Working memory | 0.52 | 6.40 | 0.00 | 0.00 |
| Total IQ | WASI verbal compre. | 0.84 | 16.23 | 0.00 | 0.00 |
| Total IQ | WASI matrix reason. | 0.77 | 12.61 | 0.00 | 0.00 |
| Total IQ | DCCS executive function. | 0.29 | 3.10 | 0.00 | 0.34 |
| Total IQ | PANSS total | -0.19 | -1.99 | 0.05 | 1.00 |
| Total IQ | PANSS positive | -0.21 | -2.20 | 0.03 | 1.00 |
| Total IQ | PANSS negative | -0.40 | -4.49 | 0.00 | 0.00 |
| Total IQ | General psychopathol. | -0.14 | -1.49 | 0.14 | 1.00 |
| Total IQ | NW18 | -0.12 | -1.29 | 0.20 | 1.00 |
| Total IQ | NW29 | -0.39 | -4.39 | 0.00 | 0.00 |
| Total IQ | NW30 | -0.39 | -4.34 | 0.00 | 0.00 |
| Picture vocab. test | Oral reading recog. | 0.69 | 10.01 | 0.00 | 0.00 |

| V1 | V2 | estimate | statistic | p.value | adj_p.value |
|---|---|---|---|---|---|
| Picture vocab. test | Auditory attention %corr | 0.41 | 4.71 | 0.00 | 0.00 |
| Picture vocab. test | Working memory | 0.46 | 5.37 | 0.00 | 0.00 |
| Picture vocab. test | WASI verbal compre. | 0.70 | 10.06 | 0.00 | 0.00 |
| Picture vocab. test | WASI matrix reason. | 0.33 | 3.58 | 0.00 | 0.07 |
| Picture vocab. test | DCCS executive function. | 0.37 | 4.09 | 0.00 | 0.01 |
| Picture vocab. test | PANSS total | -0.26 | -2.83 | 0.01 | 0.76 |
| Picture vocab. test | PANSS positive | -0.16 | -1.63 | 0.11 | 1.00 |
| Picture vocab. test | PANSS negative | -0.37 | -4.10 | 0.00 | 0.01 |
| Picture vocab. test | General psychopathol. | -0.15 | -1.60 | 0.11 | 1.00 |
| Picture vocab. test | NW18 | -0.27 | -2.88 | 0.00 | 0.65 |
| Picture vocab. test | NW29 | -0.32 | -3.48 | 0.00 | 0.10 |
| Picture vocab. test | NW30 | -0.43 | -4.90 | 0.00 | 0.00 |
| Oral reading recog. | Auditory attention %corr | 0.48 | 5.73 | 0.00 | 0.00 |
| Oral reading recog. | Working memory | 0.54 | 6.75 | 0.00 | 0.00 |
| Oral reading recog. | WASI verbal compre. | 0.60 | 7.70 | 0.00 | 0.00 |
| Oral reading recog. | WASI matrix reason. | 0.31 | 3.43 | 0.00 | 0.12 |
| Oral reading recog. | DCCS executive function. | 0.26 | 2.80 | 0.01 | 0.82 |
| Oral reading recog. | PANSS total | -0.27 | -2.93 | 0.00 | 0.56 |
| Oral reading recog. | PANSS positive | -0.08 | -0.87 | 0.39 | 1.00 |
| Oral reading recog. | PANSS negative | -0.36 | -3.98 | 0.00 | 0.02 |
| Oral reading recog. | General psychopathol. | -0.19 | -2.01 | 0.05 | 1.00 |
| Oral reading recog. | NW18 | -0.12 | -1.25 | 0.21 | 1.00 |
| Oral reading recog. | NW29 | -0.32 | -3.45 | 0.00 | 0.11 |
| Oral reading recog. | NW30 | -0.31 | -3.41 | 0.00 | 0.13 |
| Auditory attention %corr | Working memory | 0.49 | 5.92 | 0.00 | 0.00 |
| Auditory attention %corr | WASI verbal compre. | 0.30 | 3.23 | 0.00 | 0.22 |
| Auditory attention %corr | WASI matrix reason. | 0.42 | 4.78 | 0.00 | 0.00 |
| Auditory attention %corr | DCCS executive function. | 0.48 | 5.73 | 0.00 | 0.00 |
| Auditory attention %corr | PANSS total | -0.15 | -1.53 | 0.13 | 1.00 |
| Auditory attention %corr | PANSS positive | -0.03 | -0.35 | 0.73 | 1.00 |
| Auditory attention %corr | PANSS negative | -0.22 | -2.38 | 0.02 | 1.00 |
| Auditory attention %corr | General psychopathol. | -0.16 | -1.64 | 0.10 | 1.00 |
| Auditory attention %corr | NW18 | -0.19 | -2.01 | 0.05 | 1.00 |
| Auditory attention %corr | NW29 | -0.32 | -3.57 | 0.00 | 0.07 |
| Auditory attention %corr | NW30 | -0.34 | -3.73 | 0.00 | 0.04 |
| Working memory | WASI verbal compre. | 0.45 | 5.29 | 0.00 | 0.00 |
| Working memory | WASI matrix reason. | 0.42 | 4.78 | 0.00 | 0.00 |
| Working memory | DCCS executive function. | 0.42 | 4.86 | 0.00 | 0.00 |
| Working memory | PANSS total | -0.24 | -2.59 | 0.01 | 1.00 |
| Working memory | PANSS positive | -0.19 | -2.03 | 0.05 | 1.00 |
| Working memory | PANSS negative | -0.23 | -2.47 | 0.01 | 1.00 |
| Working memory | General psychopathol. | -0.24 | -2.52 | 0.01 | 1.00 |
| Working memory | NW18 | -0.14 | -1.44 | 0.15 | 1.00 |
| Working memory | NW29 | -0.34 | -3.79 | 0.00 | 0.03 |
| Working memory | NW30 | -0.24 | -2.58 | 0.01 | 1.00 |
| WASI verbal compre. | WASI matrix reason. | 0.38 | 4.30 | 0.00 | 0.01 |
| WASI verbal compre. | DCCS executive function. | 0.24 | 2.52 | 0.01 | 1.00 |

| V1 | V2 | estimate | statistic | p.value | adj_p.value |
|---|---|---|---|---|---|
| WASI verbal compre. | PANSS total | -0.24 | -2.60 | 0.01 | 1.00 |
| WASI verbal compre. | PANSS positive | -0.17 | -1.83 | 0.07 | 1.00 |
| WASI verbal compre. | PANSS negative | -0.40 | -4.59 | 0.00 | 0.00 |
| WASI verbal compre. | General psychopathol. | -0.14 | -1.48 | 0.14 | 1.00 |
| WASI verbal compre. | NW18 | -0.17 | -1.83 | 0.07 | 1.00 |
| WASI verbal compre. | NW29 | -0.26 | -2.77 | 0.01 | 0.89 |
| WASI verbal compre. | NW30 | -0.35 | -3.89 | 0.00 | 0.02 |
| WASI matrix reason. | DCCS executive function. | 0.29 | 3.13 | 0.00 | 0.31 |
| WASI matrix reason. | PANSS total | -0.05 | -0.48 | 0.63 | 1.00 |
| WASI matrix reason. | PANSS positive | -0.09 | -0.98 | 0.33 | 1.00 |
| WASI matrix reason. | PANSS negative | -0.23 | -2.44 | 0.02 | 1.00 |
| WASI matrix reason. | General psychopathol. | -0.12 | -1.27 | 0.21 | 1.00 |
| WASI matrix reason. | NW18 | -0.06 | -0.68 | 0.50 | 1.00 |
| WASI matrix reason. | NW29 | -0.37 | -4.11 | 0.00 | 0.01 |
| WASI matrix reason. | NW30 | -0.34 | -3.72 | 0.00 | 0.04 |
| DCCS executive function. | PANSS total | -0.08 | -0.82 | 0.42 | 1.00 |
| DCCS executive function. | PANSS positive | -0.14 | -1.45 | 0.15 | 1.00 |
| DCCS executive function. | PANSS negative | -0.08 | -0.88 | 0.38 | 1.00 |
| DCCS executive function. | General psychopathol. | -0.17 | -1.80 | 0.07 | 1.00 |
| DCCS executive function. | NW18 | -0.08 | -0.87 | 0.39 | 1.00 |
| DCCS executive function. | NW29 | -0.05 | -0.54 | 0.59 | 1.00 |
| DCCS executive function. | NW30 | -0.15 | -1.57 | 0.12 | 1.00 |
| PANSS total | PANSS positive | 0.38 | 4.24 | 0.00 | 0.01 |
| PANSS total | PANSS negative | 0.59 | 7.60 | 0.00 | 0.00 |
| PANSS total | General psychopathol. | 0.71 | 10.45 | 0.00 | 0.00 |
| PANSS total | NW18 | 0.16 | 1.67 | 0.10 | 1.00 |
| PANSS total | NW29 | 0.19 | 2.04 | 0.04 | 1.00 |
| PANSS total | NW30 | 0.13 | 1.39 | 0.17 | 1.00 |
| PANSS positive | PANSS negative | 0.14 | 1.48 | 0.14 | 1.00 |
| PANSS positive | General psychopathol. | 0.43 | 4.93 | 0.00 | 0.00 |
| PANSS positive | NW18 | 0.13 | 1.38 | 0.17 | 1.00 |
| PANSS positive | NW29 | 0.20 | 2.09 | 0.04 | 1.00 |
| PANSS positive | NW30 | 0.13 | 1.35 | 0.18 | 1.00 |
| PANSS negative | General psychopathol. | 0.38 | 4.31 | 0.00 | 0.00 |
| PANSS negative | NW18 | 0.01 | 0.15 | 0.88 | 1.00 |
| PANSS negative | NW29 | 0.02 | 0.24 | 0.81 | 1.00 |
| PANSS negative | NW30 | 0.13 | 1.34 | 0.18 | 1.00 |
| General psychopathol. | NW18 | 0.05 | 0.53 | 0.60 | 1.00 |
| General psychopathol. | NW29 | 0.18 | 1.87 | 0.06 | 1.00 |
| General psychopathol. | NW30 | 0.15 | 1.55 | 0.12 | 1.00 |
| NW18 | NW29 | 0.46 | 5.43 | 0.00 | 0.00 |
| NW18 | NW30 | 0.55 | 6.93 | 0.00 | 0.00 |
| NW29 | NW30 | 0.59 | 7.67 | 0.00 | 0.00 |
